# Immune Training with BCG therapy in Diabetes: Outcomes of a Phase 2 randomized clinical trial

**DOI:** 10.64898/2026.10.09.26365133

**Authors:** Denise L. Faustman, Joan E. Braley, Neeshi Hullavarad, Emily R. Bulczynski, Jack L. Dorsey, Nicholas G. Kartsounis, Lisa T. Tran, Bethany J. Thach, Hikaru Hayashi, Daniel A. Mounier, Lindsey A. Adams, Serena E. Bradley, Shiho Hashiguchi, Hui Zheng, Willem M. Kühtreiber

## Abstract

BCG is a well recognized old generic drug that can mold the human immune system through Immune Training. Broad basic science studies support Immune Training, but formal randomized placebo control trials are frequently lacking for confirmation of human benefits. Here we report on a randomized, double-blind, placebo-controlled phase 2 trial with up to 8 years of follow-up. The primary outcome was mean percent change from baseline in HbA1c. At year 5, HbA1c in the BCG group was reduced from baseline by 7.04% (P=0.0006) and at year 8 by 12.84% (P=0.0001), whereas HbA1c did not change from baseline in the placebo group at either time point. Longitudinal percentage changes in HbA1c from baseline using mixed model repeated measures analysis with a comparison of BCG versus Placebo arms was significant (p=0.0017). This shows durable long term improvements lowering of HbA1c only with BCG immunotherapy. Secondary outcomes were also significant. There was a statistically significant reduction in average insulin dose only in the BCG-treated group (P=0.024). Glycemic improvements occurred without recovery of C-peptide from negligible baseline levels, suggesting no pancreas contribution. Blood sugar values obtained from continuous glucose monitoring (CGM) similarly showed BCG immunotherapy lowers blood sugars compared to the Placebo arm (P=0.0189). CGM data also showed BCG immunotherapy resulted in an up to 183.7% improvement over baseline in time spent in normoglycemia (blood sugar in the normal range of 70-99 mg/dL) (P=0.0391). A sensitivity analysis confirmed BCG specific reductions in HbA1c were robust and durable at least to Year 08 (*P<0.0001*). BCG was safe. These finding support improved blood sugar control through immune training of metabolism without a contribution of the pancreas even after long standing T1D. Trial registration: NCT02081326.

## Introduction

Type 1 diabetes (T1D) is an autoimmune disease marked by progressive loss of pancreatic beta-cell function, resulting in a rapid decline in endogenous insulin secretion and lifelong dependence on exogenous insulin. Despite significant advances in diabetes management, major challenges persist. About 75% of patients do not achieve recommended glycemic targets^1^; exogenous insulin remains an imperfect substitute for the physiologic function of pancreatic beta cells; hypoglycemia is common; and diabetes-related complications continue to occur. Further, the daily burden of self-management is substantial, and the costs of treatment remain high, roughly 2.6 times higher than that for people without diabetes^2^. Together, these limitations underscore the need for safe, more effective, less burdensome, and more affordable therapies to control blood sugars.

Mycobacteria are ancient organisms that co-evolved with humans and helped shape their immune system regulation^3^. Bacille-Calmette-Guérin (BCG), a live, avirulent strain of *Mycobacterium bovis,* was first introduced more than 100 years ago as a vaccine to prevent human tuberculosis (TB). BCG has been in continuous use since then, and today is administered to nearly 90% of newborns worldwide^4^. BCG is closely related to pathogenic *Mycobacterium tuberculosis*, the causative agent of human TB. Both BCG and *Mycobacterium tuberculosis* have been shown to stably induce immune system regulatory T cells (Tregs)^5–9^, which suppress immune responses through immune training and may be beneficial for curtailing autoimmunity. Over the last 15 years, clinical trials have evaluated BCG as investigational immunotherapy in at least two autoimmune diseases, multiple sclerosis^10–12^ and autoimmune diabetes^13–19^.

In our earlier small phase 1A randomized clinical trial in adults with longstanding T1D^17^, two doses of BCG showed desirable acute effects on innate immune training through release of tumor necrosis factor, which causes selective death of insulin-autoreactive T cells^20–22^. But the trial’s duration of only 20 weeks was, in retrospect, too short to have observed an improvement in hemoglobin A1c (HbA1c) with a durable effect on the autoimmune process^17^. FDA draft guidance considers HbA1c reduction the primary outcome for evaluating efficacy of new anti-diabetic drugs or biologics for glucose control^23^. When we extended follow-up of our phase 1A trial from 20 weeks to 8 years in a Phase 1B trial, BCG was shown to stably and durably lower HbA1c to near-normal levels, after a time lag of approximately 2-3 years^14^.

The lowering of HbA1c without a functioning pancreas was found in our earlier studies to be due to correction of an underlying T1D defect in the lymphoid (and myeloid) system, namely, an overreliance of energy metabolism on oxidative phosphorylation, an energy generating step that typically uses fats as the starting source^14–16,18,19,24–26^. BCG immunotherapy slowly corrected this underlying defect in T1D lymphoid (and myeloid) cellular metabolism by conversion to aerobic glycolysis, a metabolic pathway marked by increased glucose utilization^16,18^. The improved glucose regulation was associated with increased glucose utilization by the spleen, the largest lymphoid organ^27^. This clinical trial example of Immune Training with the passage of time from the introduction of BCG was due to epigenetic reset of many lymphoid and myeloid cell genes toward aerobic glycolysis pathways^24,25^. There is now a large literature of how bacterial organisms or vaccines themselves can reprogram the human host and have major impacts on host gene methylation patterns^28–45^

The current study is a phase 2 randomized, double-blind, placebo-controlled trial of multi-dose BCG in adults with longstanding T1D. Here we report on 5-year and 8-year outcomes of a 10-year trial. The prespecified primary outcome is mean % change from baseline in HbA1c. Secondary outcomes include insulin dose, time spent in normoglycemia, IDAA1c and other continuous glucose monitoring outcomes, autoantibody levels, and stimulated C-peptide secretion.

## Results

### Baseline

Baseline characteristics of T1D participants appear in Table 1, and participant flow is shown in the CONSORT diagram (Fig. 1). At trial start, T1D had a mean disease duration of 14.5 ± 1.5 years. Subjects were recruited from across the United States, and their diabetes care was driven by their primary endocrinologist (Fig. S1). Overall, 53 out of 58 enrolled patients completed the trial at 5-years, the first time point for data analysis (retention rate 91.4%; see Fig. S2). The second data analysis time point is 8-years. This study has a total duration of 10 years since trial to prove BCG immunotherapy durability. This trial also has an option at 5-years of participating in the cross-over study (data not shown).

**Fig. 1.**
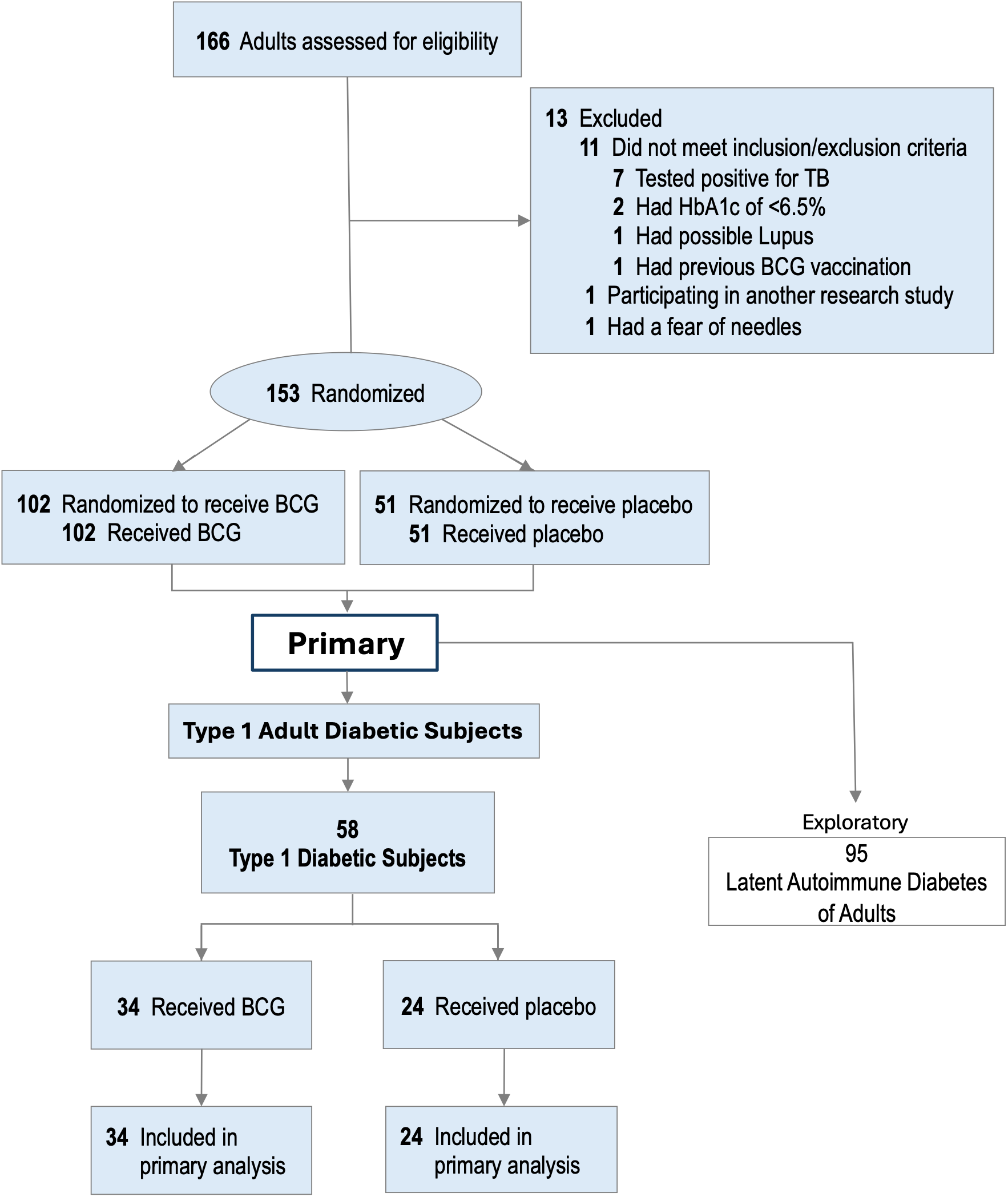
Consort Flow Diagram of Adult Participants in a Five-Year Trial of BCG Immunotherapy on Longstanding Type 1 Diabetes

**Table 1.**
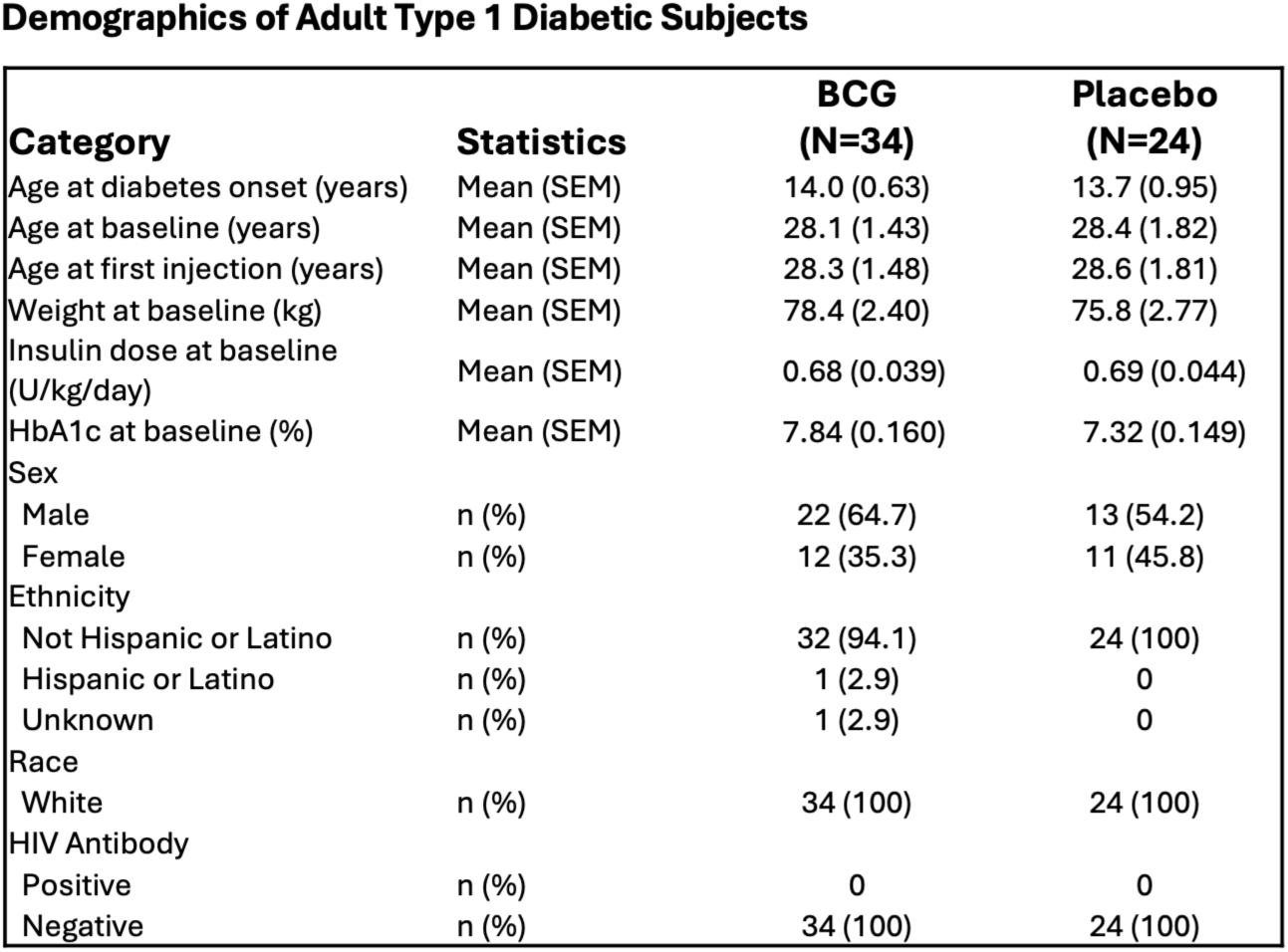

| **Category** | **Statistics** | **BCG**<br>**(N=34)** | **Placebo**<br>**(N=24)** |
| --- | --- | --- | --- |
| Age at diabetes onset (years) | Mean (SEM) | 14.0 (0.63) | 13.7 (0.95) |
| Age at baseline (years) | Mean (SEM) | 28.1 (1.43) | 28.4 (1.82) |
| Age at first injection (years) | Mean (SEM) | 28.3 (1.48) | 28.6 (1.81) |
| Weight at baseline (kg) | Mean (SEM) | 78.4 (2.40) | 75.8 (2.77) |
| Insulin dose at baseline<br>(U/kg/day) | Mean (SEM) | 0.68 (0.039) | 0.69 (0.044) |
| HbA1c at baseline (%) | Mean (SEM) | 7.84 (0.160) | 7.32 (0.149) |
| Sex |  |  |  |
| Male | n (%) | 22 (64.7) | 13 (54.2) |
| Female | n (%) | 12 (35.3) | 11 (45.8) |
| Ethnicity |  |  |  |
| Not Hispanic or Latino | n (%) | 32 (94.1) | 24 (100) |
| Hispanic or Latino | n (%) | 1 (2.9) | 0 |
| Unknown | n (%) | 1 (2.9) | 0 |
| Race |  |  |  |
| White | n (%) | 34 (100) | 24 (100) |
| HIV Antibody |  |  |  |
| Positive | n (%) | 0 | 0 |
| Negative | n (%) | 34 (100) | 24 (100) |

### Primary outcome: % change from baseline in HbA1c

The BCG arm had a significant mean reduction in HbA1c from baseline of 7.04 ± 2.69% (mean ± SEM; *P* = 0.0006) at year 5 and 12.84 ± 2.71% (*P* = 0.0001) at year 8 (Fig. 2A and Table 2). The placebo group had no significant % change from baseline in HbA1c at either year 5 or year 8 (P=0.0781 at year 5; *P=0.500* at year 8). The BCG (vs placebo) group showed greater % reduction with Mixed Model Repeated Measures comparison of BCG versus placebo arms (*P=0.0017*) (Fig. 2A and Table 2). All data is represented as Mean ± SEM (Table 2A) and as LS Mean and 95%CI (Table 2B).

**Fig. 2.**
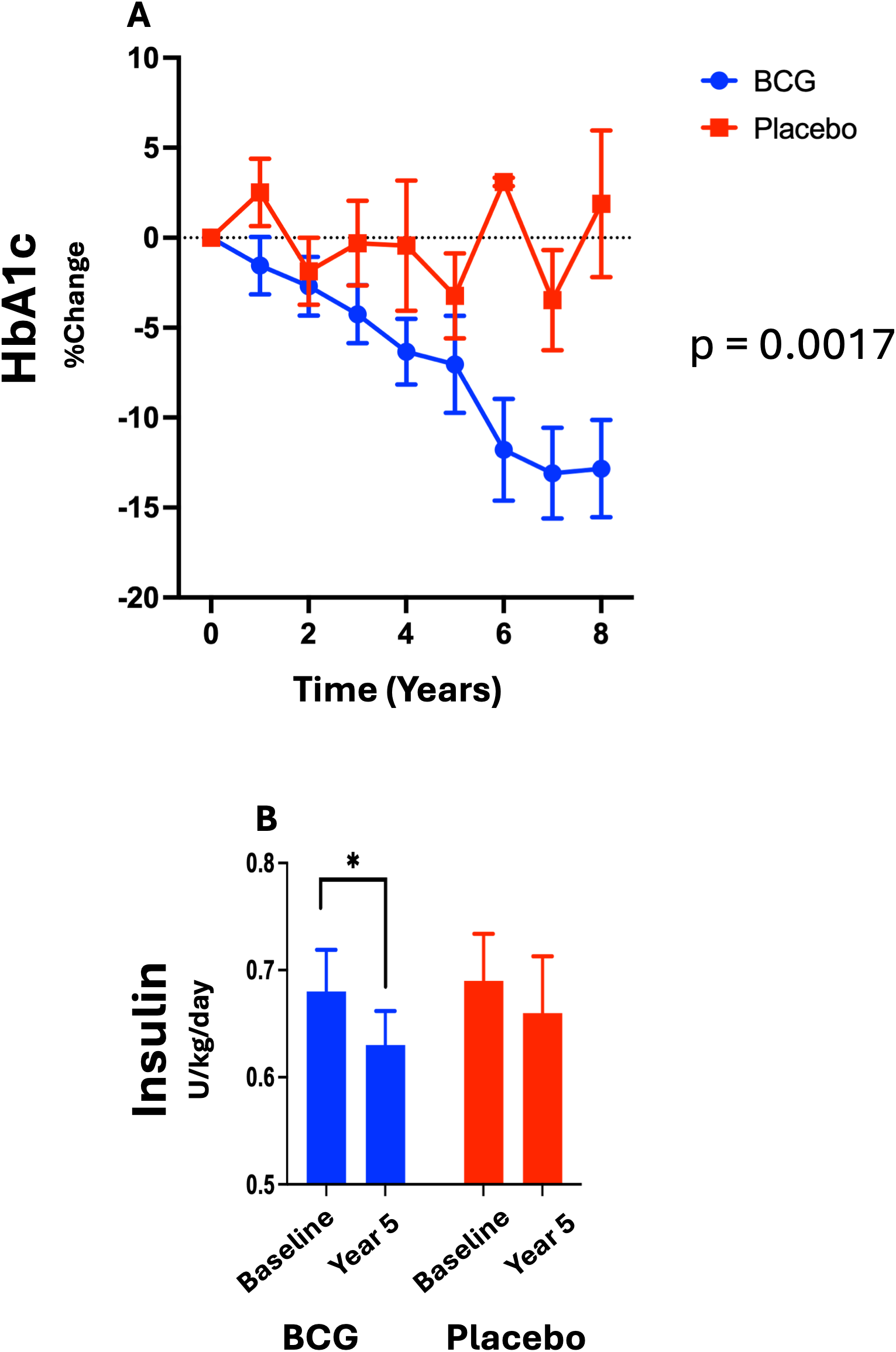
Efficacy of Multi-Dose BCG Immunotherapy: HbA1c and Insulin Use **(A) Effect of BCG immunotherapy on HbA1c**. As compared to baseline, at Year 5 the BCG arm had a reduction in HbA1c of 7.04 ± 2.695% (Mean ± SEM, Wilcoxon Signed Rank Test one-tailed P=0.0006). By Year 8 the reduction had reached 12.84 ± 2.705% (P=0.0001). In the placebo group there was no change (Year 5 P=0.0781 and Year 8 P=0.500). BCG n=34, placebo n=24. **(B) Effect of BCG immunotherapy on insulin use** Fasting insulin dose (Units/kg/day) in BCG-treated and placebo-treated adult subjects at Year 0 vs Year 5. In the BCG-treated group, insulin use decreased from 0.68 ± 0.04 U/kg/day at Year 0 to 0.63 ± 0.03 by Year 5 (Mean ± SEM, *P=*0.024). There was no significant reduction in average insulin use in the placebo group *(P=*0.160) (Paired Student’s t-test, one-tailed). * *P <0.05* BCG n=34, placebo n=24.

**Table 2.**
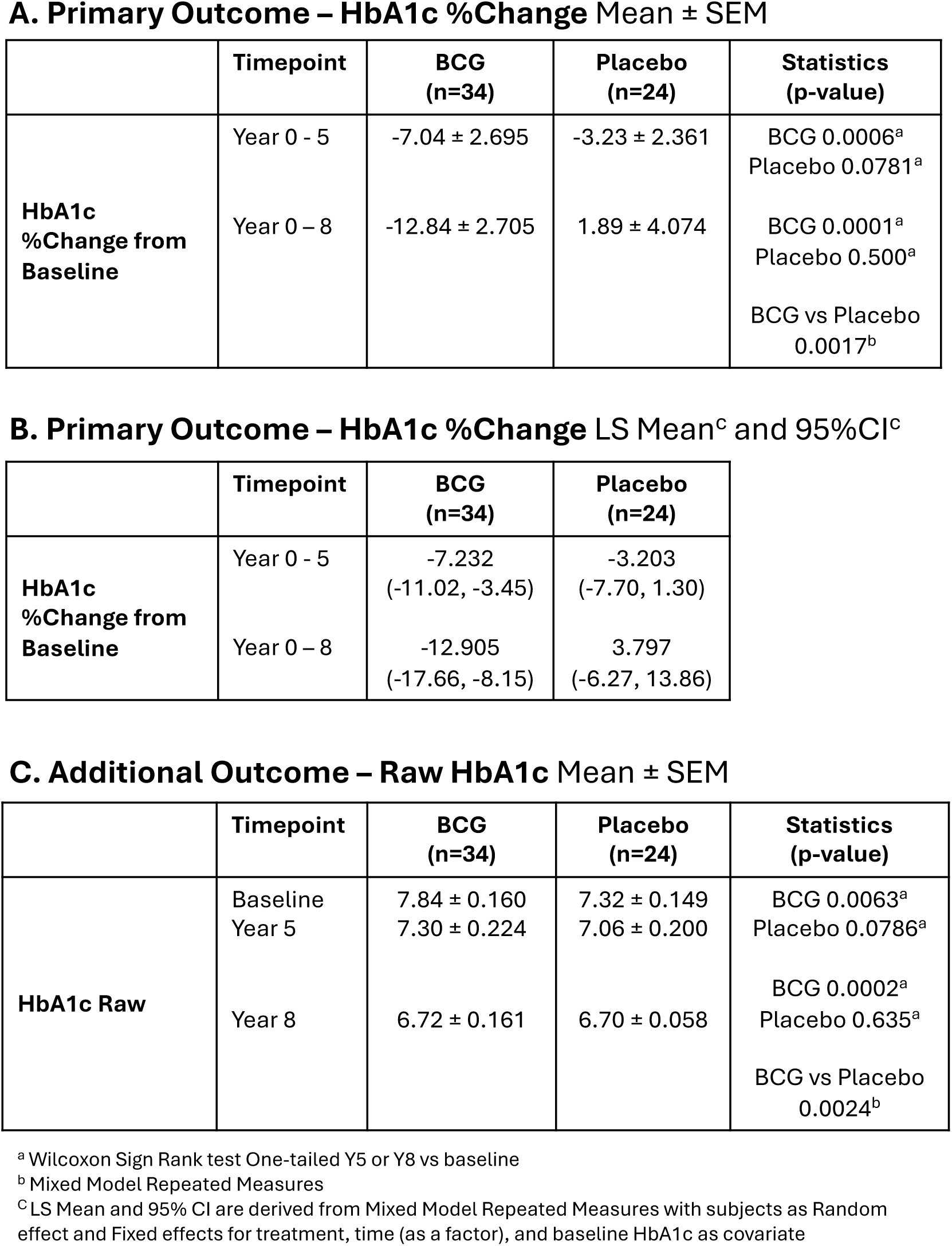

Although not an outcome, a supportive analysis of raw HbA1c in the BCG group shows similar strong statistical significance in the BCG group and no significance in the placebo group (Table 2C). As demonstrated previously from in vitro studies^15^ the BCG-related drop in HbA1c in this trial was greatest for adults with earlier AOO (age of onset) (Fig. 3). The largest average BCG-related drop in HbA1c at Year 5 was -18.89 ± 3.63% for subjects with AOO <12 years old (BCG vs placebo at Year 5 (Mixed Model Repeated Measures *P=0.009*). Also BCG treated T1D with AOO <13 and < 16 both had a statistically significant drop in HbA1c compared to the placebo group (Mixed Model Repeat Measurements, *P=0.031, P=0.03*). There was no impact of disease duration on the magnitude of HbA1c drop (data not shown).

**Fig. 3.**
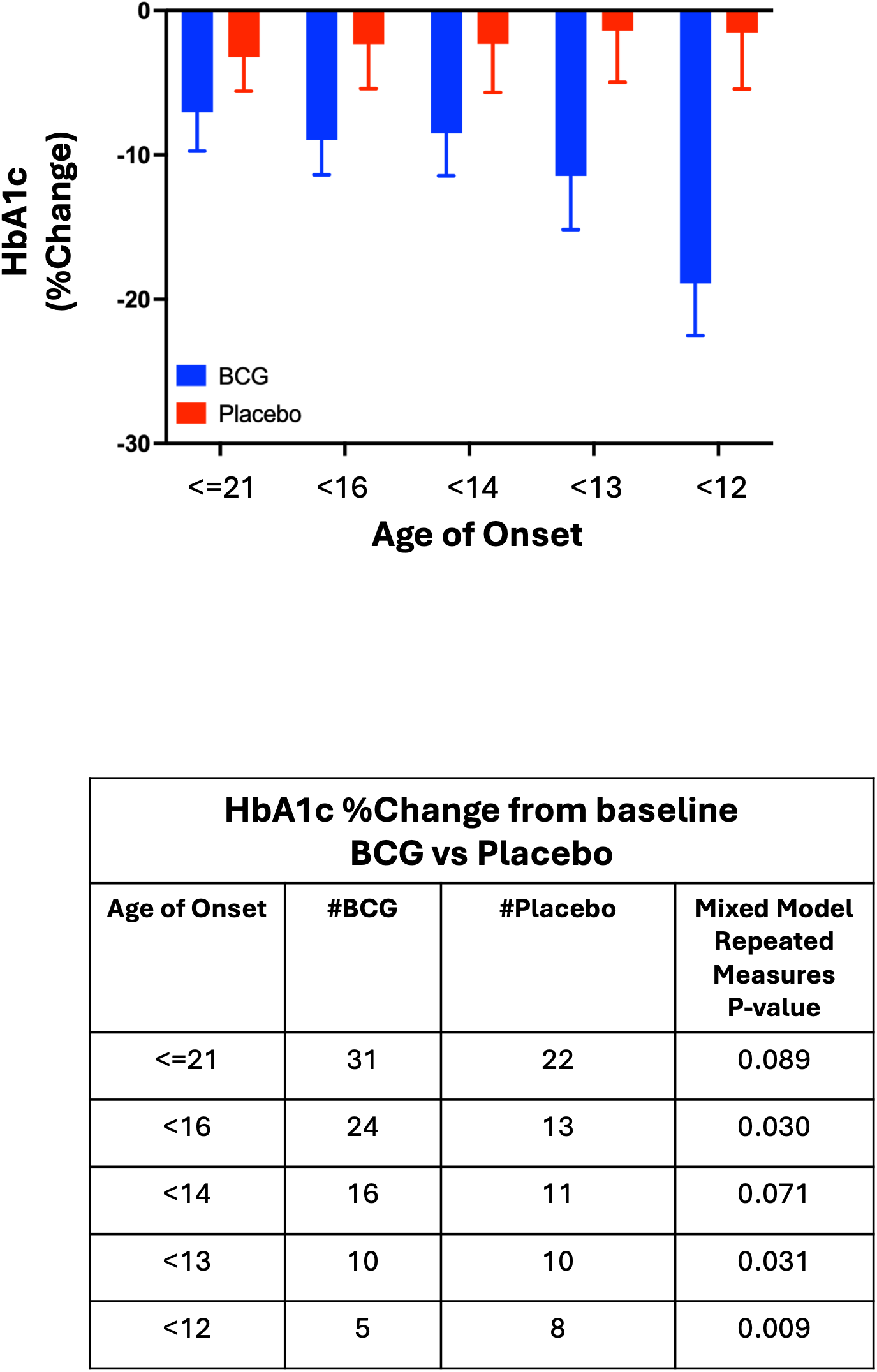
Efficacy of BCG is correlated with Age of Onset HbA1c lowering calculated as a % change based on age of onset, highlights the magnitude of the HbA1c lowering based on age of onset. The figure shows HbA1c data expressed as the % change from Year 0 versus Year 5. The data show the greatest drop of HbA1c occurs in the BCG-treated group with age of onset less than 12 years of age with a -18.89 ± 3.63% (Mean ± SEM) drop. For the placebo group, the HbA1c level remained essentially unchanged at -1.52 ± 3.91%. The difference between BCG and placebo groups at the AOO <12, AOO<13 and AOO<16 groups were significant (Mixed Model Repeated Measures).

### Secondary outcome: insulin dose

Mean insulin use in the BCG group decreased from 0.68 to 0.63 U kg⁻¹ day⁻¹ from baseline to year 5 (*P = 0.024;* Fig. 2b). There was no significant reduction over time in the placebo group *(P=0.160*). The significant BCG-related % drop in HbA1c described above occurred *despite* a reduction in insulin dose, ruling out a change in HbA1c from exogenous therapy as the cause for the drop.

We also calculated IDAA1c from insulin dose (Fig 2B) and body weight (Supplemental Fig 3) and HbA1c to ascertain that the above reduction in HbA1c was not due to some combination of changes in insulin use and body weight. Comparison of BCG and Placebo arms for IDAA1c indeed showed a continuing significant difference (Mann-Whitney BCG vs Placebo p=0.047). Therefore the drop in HbA1c with BCG immunotherapy in T1D was not due to more insulin, not due to weight loss and was not due to the standard of care changing.

### CGM measures of glycemic control

Values obtained from continuous glucose monitoring (CGM) showed lowering of average blood sugars over the course of the trial in the BCG arm as compared to the placebo arm (*P=0.0189;* Fig. 4A). CGM data also showed that BCG immunotherapy was associated with up to 183.7% improvement over baseline in time spent in normoglycemia (blood sugar in the normal range of 70-99 mg/dL) (Fig. 4Bi; *P=0.0391*). This improvement was not observed in the placebo group (*P=0.5625*). Despite the significant increase in normoglycemia in the BCG arm over time, the comparison between BCG and placebo arms was not significant (Mixed Model Repeated Measures *P=0.0849*). BCG immunotherapy was associated with less hyperglycemia as compared to placebo control (Mixed Model Repeated Measures *P=0.0110*; Fig. 4Bii).

**Fig. 4.**
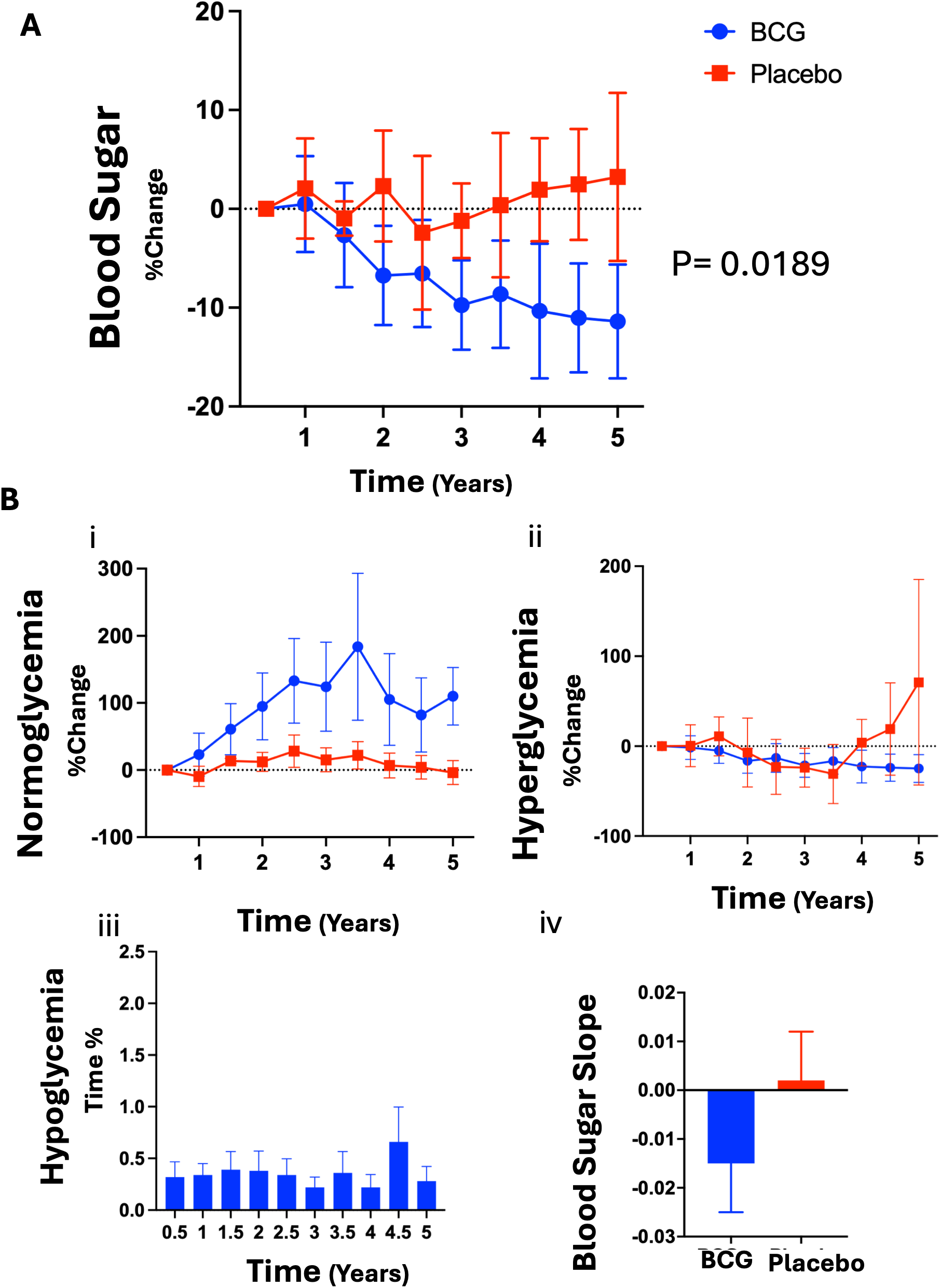
CGM Monitoring: Average Blood Sugar, Time in Normoglycemia, Hyperglycemia, and Hypoglycemia. **(A)** CGM data shows that the average blood sugar decreased in the BCG group, but not in the placebo group. Longitudinal CGM data was averaged over periods of 6 months each and then graphed as the % change from baseline. Over 5 years of follow-up, the BCG group showed a reduction in average blood sugar of 11.39 ± 5.764% (Mean ± SEM) as compared to baseline, whereas the placebo group showed an increase of 3.23 ± 8.511%. Mixed Model Repeated Measures analysis showed that the difference between BCG and placebo was significant (P=0.0189). **(Bi)** The data represent the percentage of continuous glucose monitoring (CGM) data points in the normoglycemic range (70-99 mg/dL) versus the total number of CGM data points. Since each CGM data point captured the same amount of time (5 minutes), the percentage CGM data in the normoglycemic range is equivalent to the percentage of time in normoglycemia for a given 24-hr period. The average percent of time with normoglycemia was calculated across all T1D patients in the BCG and placebo group separately, and then expressed as the change from baseline (Year 0) over the 5-year trial. While time in normoglycemic range remained essentially unchanged in the placebo group (p=0.5625), the BCG group increased normoglycemia by up to nearly 200% compared to baseline (p=0.0391). Comparison of BCG versus placebo arms was a trend (Mixed Model Repeated Measures p=0.0849). **(Bii)** Dexcom CGM data for all T1D participants was used to determine for each subject and for each semi-annual time period the percentage of CGM data points in the hyperglycemic range (>180 mg/dL) versus the total number of CGM data points. Since each CGM data point represents the same amount of time (5 minutes), this was equivalent to the percentage of hyperglycemia time. While the subjects in the placebo group spent increasing amounts of time in the hyperglycemic range over the course of the trial, there was a steady decrease in hyperglycemia time in the BCG group although the difference was a desirable trend (BCG vs placebo; *P*=0.083; Linear Mixed Effect Modeling). **(Biii)** The number of hypoglycemic CGM data points (blood sugar <45 mg/dL) for each BCG treated patient in each semiannual period was tabulated and the result was expressed as a percentage of the total number of CGM data points. Calculations show the averages and SEM and plotted over time. The hypoglycemia rate remained unchanged across the 5-year trial period in the BCG group as the HbA1c and average blood sugar fell over the same time period. **(Biv)** The slope of mean glucose levels over 5-years of follow-up. This was a linear regression analysis through all CGM data points over time for each patient separately and then an average of the slopes for all patients in the BCG and placebo groups. The average slope in the BCG group was negative, indicating that, overall, the blood glucose in BCG-treated subjects decreased over time, whereas it remained unchanged or increased slightly in the placebo group. The difference was significant. (* *P*=0.036; Mann-Whitney, one-tailed)

With the BCG immunotherapy-related improvements in HbA1c, IDAA1c, time in normoglycemia and prevention of hyperglycemia, it was important that incidence of hypoglycemia (blood sugar <45 mg/dL) did not increase with BCG immunotherapy. This data is shown in Fig. 4Biii, and suggests that BCG immunotherapy is associated with restored glucose control without increasing hypoglycemia.

The linear regression slope of blood sugars, calculated per subject over all CGM data by 5-year follow-up and then averaged, also decreased with BCG over the study period of 5 years, while the slope with placebo remained unchanged (BCG versus placebo *P=0.036*; Fig. 4Biv).

### Pancreas activity

Activity of pancreatic islet β-cells is captured by fasting or stimulated C-peptide output, with stimulated C-peptide providing an accurate measure of pancreas insulin reserve. At yearly intervals all clinical trial subjects received an IV glucagon challenge test to assess pancreas reserve. In adults with T1D and a disease duration averaging 14.5 ± 1.5-years, fasting C-peptide levels were very low at randomization, suggesting that many of these subjects, as expected with long term diabetes, had no detectable C-peptide on their first treatment visit to the clinic (Fig 5A). The data showed that 48.3% of the patients had undetectable (≤5 pmol/L) fasting levels of C-peptide at the trial’s start; the average fasting mean C-peptide level was 21.88 ± 3.61 pmol/L for the group overall. At year 5 BCG treatment did not restore C-peptide after this state of near absence (Fig. 5B). There was no significant difference between the BCG and placebo (Mann-Whitney at Year 5 *P=0.5827*; Mixed Model Repeated Measures *P=0.2553*). This implies that BCG’s therapeutic effect of blood sugar lowering is pancreas independent, and is likely a correction of an underlying metabolic defect in the lymphoid system.

**Fig. 5.**
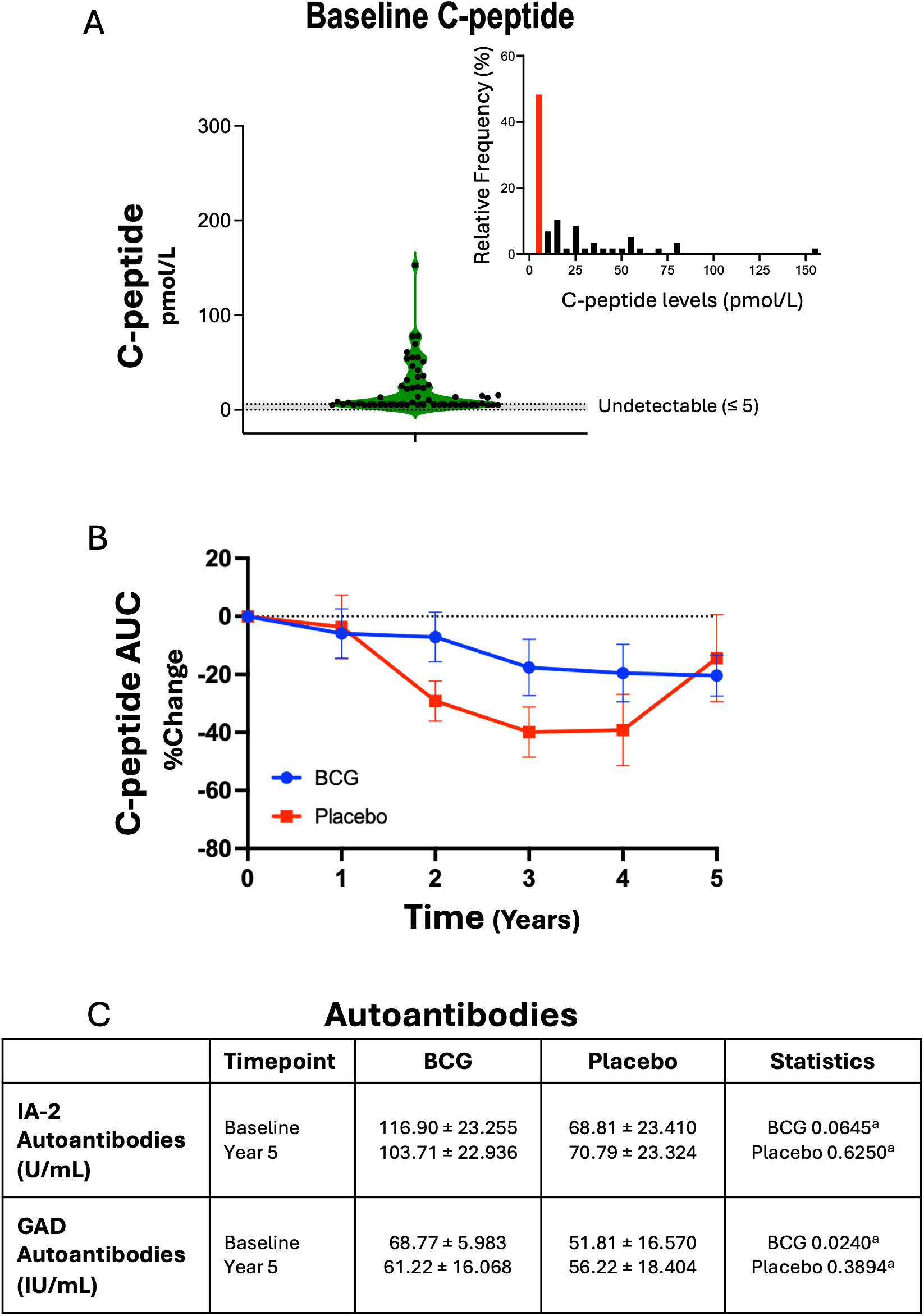
Effect of BCG immunotherapy on endogenous C-peptide secretion **(A)** Distribution of fasting C-peptide serum levels at baseline (Year 0). The violin plot shows that 48.3% of the patients had undetectable fasting C-peptide at the first BCG vaccine visit, even for detection of very low levels (≤5 pmol/L). The inset shows a histogram of the distribution of fasting C-peptide for all enrolled T1D subjects. The red bar in the histogram shows that 48.3% of all T1D subjects at the start of the trial have undetectable fasting C-peptide serum levels. The average C-peptide was 21.88 ± 3.61 pmol/L (Mean ± SEM). BCG n=34, placebo n=24. **(B)** Analysis of the area under the curve (AUC) for C-peptide during the GST (Glucagon Stimulation Test) performed at each annual visit for 5 years. C-peptide secretory capacity using a yearly Glucagon Simulation Test (GST) was used as a measure of pancreas insulin reserve. ELISA was used to quantify C-peptide in each serum sample collected during the GST. Using the trapezoid method, for each subject we calculated the area under the curve (AUC) for each of their annual GSTs. We then calculated the percent change from baseline (Year 0) versus each subsequent yearly GST and averaged across all subjects in the BCG and placebo groups separately. The subjects in both BCG and placebo groups showed a gradual decrease in their capacity to respond to the glucagon bolus with the secretion of C-peptide. There was no significant difference between the BCG and the placebo *(P=*0.5827; Mann-Whitney, one-tail at Year 5; Mixed Model Repeated Measures P=0.2553). BCG n=34, placebo n=24. (C) Analysis of autoantibodies IA-2 and GAD Levels of antibodies against IA-2 and GAD measured using ELISA at baseline and Year 5. For IA-2 autoantibodies, there was a downward trend within the BCG treatment arm (p=0.0645), but not in the placebo arm (p=0.6250). For GAD autoantibodies, there was a significant decrease in the BCG arm (p=0.0240), but not in the placebo arm (p=0.3894). BCG n=34, placebo n=24.

### Autoantibodies

BCG has been shown to induce Treg cells, but in this Phase 2 trial of long term T1Ds, there was little or no pancreas remaining to preserve^46–48^. To understand if there was any underlying change in the persistent primary autoimmune disease, five-year long monitoring of diabetes related autoantibodies was studied. The data in Fig 5C shows that there was a significant decrease from baseline in GAD autoantibodies in the BCG arm (*P=0.0240*), but not in the placebo arm (*P=0.3894*). For IA-2 autoantibodies, there was a trend for a decrease in the BCG arm (*P=0.0645*), but not in the placebo arm (*P=0.6250*). Taken together, this data suggests that BCG treatment appears to be decreasing autoimmunity, at least assessed in terms of reduction in two known autoantibodies associated with T1D.

### Sensitivity analysis

A sensitivity analysis was performed to look for differences in HbA1c from baseline to year 8 by calculating the P-value obtained from the average value of 100 imputations based on the LSM estimate in a Mixed Model for Repeated Measures (MMRM) analysis. The model included treatment and time (as a categorical factor) as fixed effects, with baseline HbA1c included as a covariate. This sensitivity analysis was highly significant by Year 8 (*P<0.0001*), confirming that the BCG-specific reductions in HbA1c were statistically robust and durable at least to year 08.

### Safety

Safety data are reported (Table 3, 4), which covers 18 visits, all fasting. The BCG vaccine can cause erythema at the injection site, 4 weeks to 2 months afterward; no vaccine erythema exceeded the expected range as documented by a third-party blinded to study group.

**Table 3.**
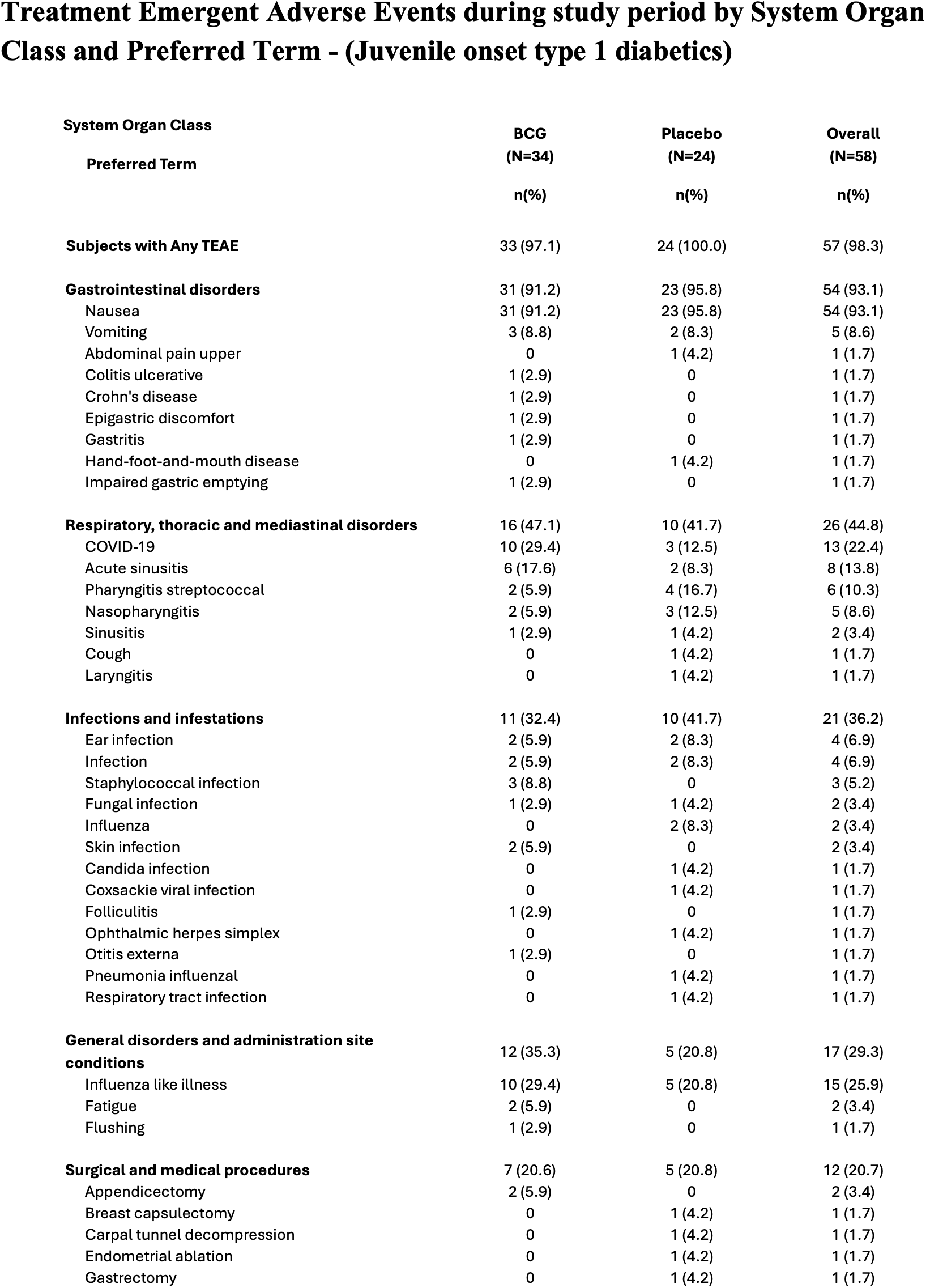
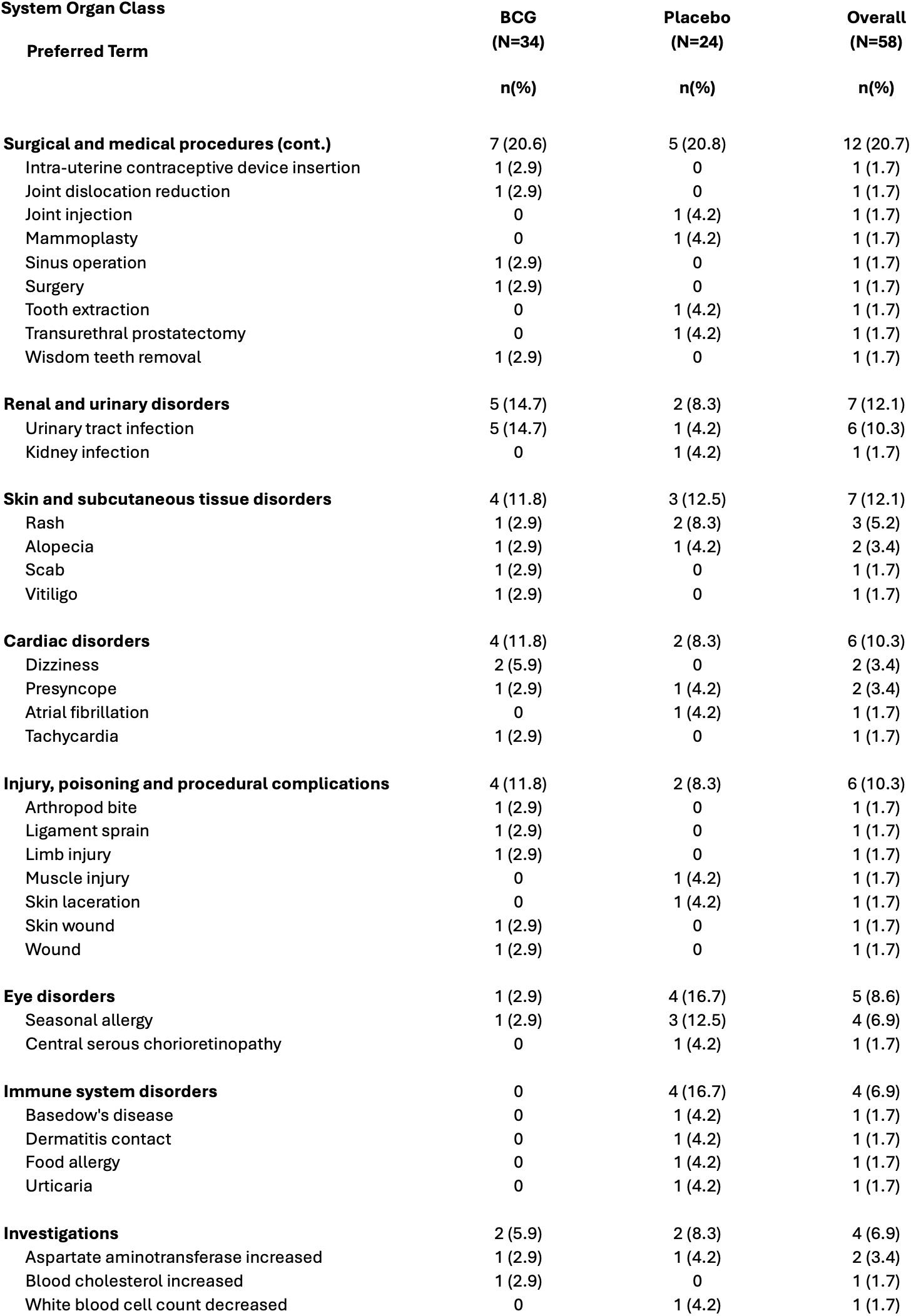
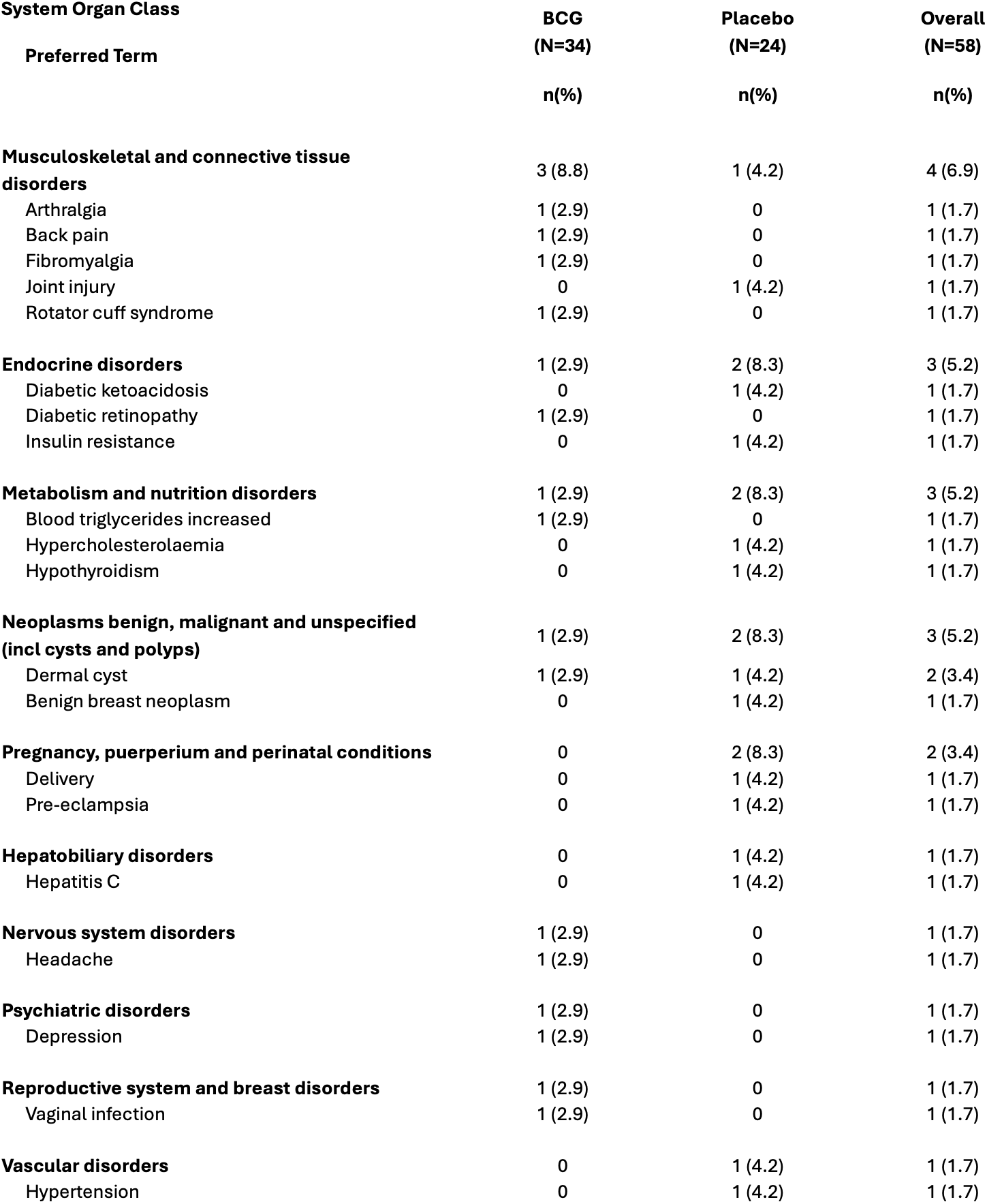
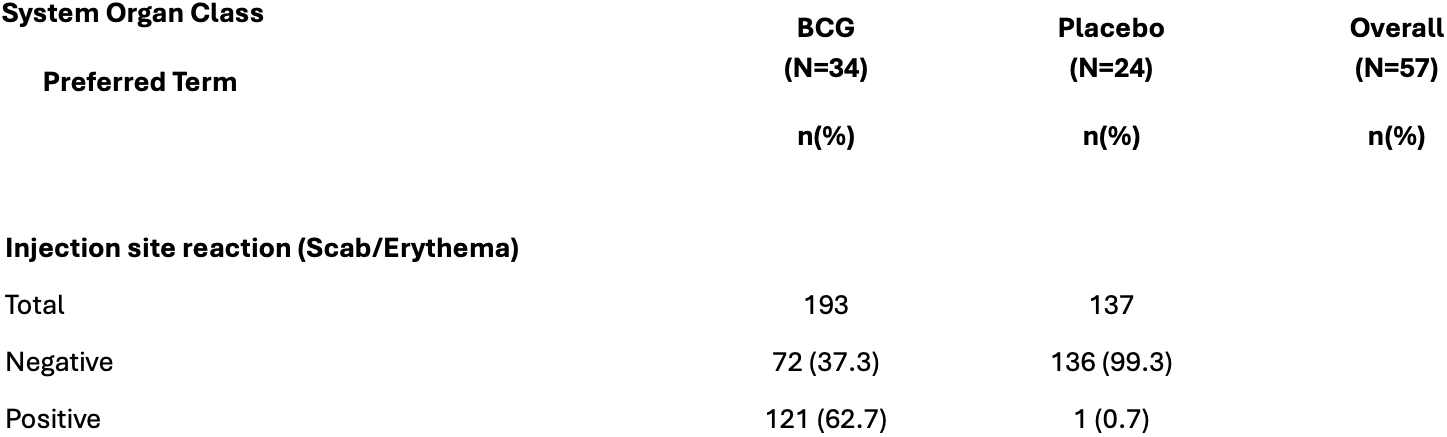

| **System Organ Class** | **BCG**<br>**(N=34)** | **Placebo**<br>**(N=24)** | **Overall**<br>**(N=58)** |
| --- | --- | --- | --- |
| **Preferred Term** | **n(%)** | **n(%)** | **n(%)** |
| **Subjects with Any TEAE** | 33 (97.1) | 24 (100.0) | 57 (98.3) |
| **Gastrointestinal disorders** | 31 (91.2) | 23 (95.8) | 54 (93.1) |
| Nausea | 31 (91.2) | 23 (95.8) | 54 (93.1) |
| Vomiting | 3 (8.8) | 2 (8.3) | 5 (8.6) |
| Abdominal pain upper | 0 | 1 (4.2) | 1 (1.7) |
| Colitis ulcerative | 1 (2.9) | 0 | 1 (1.7) |
| Crohn's disease | 1 (2.9) | 0 | 1 (1.7) |
| Epigastric discomfort | 1 (2.9) | 0 | 1 (1.7) |
| Gastritis | 1 (2.9) | 0 | 1 (1.7) |
| Hand-foot-and-mouth disease | 0 | 1 (4.2) | 1 (1.7) |
| Impaired gastric emptying | 1 (2.9) | 0 | 1 (1.7) |
| **Respiratory, thoracic and mediastinal disorders** | 16 (47.1) | 10 (41.7) | 26 (44.8) |
| COVID-19 | 10 (29.4) | 3 (12.5) | 13 (22.4) |
| Acute sinusitis | 6 (17.6) | 2 (8.3) | 8 (13.8) |
| Pharyngitis streptococcal | 2 (5.9) | 4 (16.7) | 6 (10.3) |
| Nasopharyngitis | 2 (5.9) | 3 (12.5) | 5 (8.6) |
| Sinusitis | 1 (2.9) | 1 (4.2) | 2 (3.4) |
| Cough | 0 | 1 (4.2) | 1 (1.7) |
| Laryngitis | 0 | 1 (4.2) | 1 (1.7) |
| **Infections and infestations** | 11 (32.4) | 10 (41.7) | 21 (36.2) |
| Ear infection | 2 (5.9) | 2 (8.3) | 4 (6.9) |
| Infection | 2 (5.9) | 2 (8.3) | 4 (6.9) |
| Staphylococcal infection | 3 (8.8) | 0 | 3 (5.2) |
| Fungal infection | 1 (2.9) | 1 (4.2) | 2 (3.4) |
| Influenza | 0 | 2 (8.3) | 2 (3.4) |
| Skin infection | 2 (5.9) | 0 | 2 (3.4) |
| Candida infection | 0 | 1 (4.2) | 1 (1.7) |
| Coxsackie viral infection | 0 | 1 (4.2) | 1 (1.7) |
| Folliculitis | 1 (2.9) | 0 | 1 (1.7) |
| Ophthalmic herpes simplex | 0 | 1 (4.2) | 1 (1.7) |
| Otitis externa | 1 (2.9) | 0 | 1 (1.7) |
| Pneumonia influenzal | 0 | 1 (4.2) | 1 (1.7) |
| Respiratory tract infection | 0 | 1 (4.2) | 1 (1.7) |
| **General disorders and administration site conditions** | 12 (35.3) | 5 (20.8) | 17 (29.3) |
| Influenza like illness | 10 (29.4) | 5 (20.8) | 15 (25.9) |
| Fatigue | 2 (5.9) | 0 | 2 (3.4) |
| Flushing | 1 (2.9) | 0 | 1 (1.7) |
| **Surgical and medical procedures** | 7 (20.6) | 5 (20.8) | 12 (20.7) |
| Appendectomy | 2 (5.9) | 0 | 2 (3.4) |
| Breast capsulectomy | 0 | 1 (4.2) | 1 (1.7) |
| Carpal tunnel decompression | 0 | 1 (4.2) | 1 (1.7) |
| Endometrial ablation | 0 | 1 (4.2) | 1 (1.7) |
| Gastrectomy | 0 | 1 (4.2) | 1 (1.7) |
| **Preferred Term** | **n(%)** | **n(%)** | **n(%)** |
| **Surgical and medical procedures (cont.)** | 7 (20.6) | 5 (20.8) | 12 (20.7) |
| Intra-uterine contraceptive device insertion | 1 (2.9) | 0 | 1 (1.7) |
| Joint dislocation reduction | 1 (2.9) | 0 | 1 (1.7) |
| Joint injection | 0 | 1 (4.2) | 1 (1.7) |
| Mammoplasty | 0 | 1 (4.2) | 1 (1.7) |
| Sinus operation | 1 (2.9) | 0 | 1 (1.7) |
| Surgery | 1 (2.9) | 0 | 1 (1.7) |
| Tooth extraction | 0 | 1 (4.2) | 1 (1.7) |
| Transurethral prostatectomy | 0 | 1 (4.2) | 1 (1.7) |
| Wisdom teeth removal | 1 (2.9) | 0 | 1 (1.7) |
| **Renal and urinary disorders** | 5 (14.7) | 2 (8.3) | 7 (12.1) |
| Urinary tract infection | 5 (14.7) | 1 (4.2) | 6 (10.3) |
| Kidney infection | 0 | 1 (4.2) | 1 (1.7) |
| **Skin and subcutaneous tissue disorders** | 4 (11.8) | 3 (12.5) | 7 (12.1) |
| Rash | 1 (2.9) | 2 (8.3) | 3 (5.2) |
| Alopecia | 1 (2.9) | 1 (4.2) | 2 (3.4) |
| Scab | 1 (2.9) | 0 | 1 (1.7) |
| Vitiligo | 1 (2.9) | 0 | 1 (1.7) |
| **Cardiac disorders** | 4 (11.8) | 2 (8.3) | 6 (10.3) |
| Dizziness | 2 (5.9) | 0 | 2 (3.4) |
| Presyncope | 1 (2.9) | 1 (4.2) | 2 (3.4) |
| Atrial fibrillation | 0 | 1 (4.2) | 1 (1.7) |
| Tachycardia | 1 (2.9) | 0 | 1 (1.7) |
| **Injury, poisoning and procedural complications** | 4 (11.8) | 2 (8.3) | 6 (10.3) |
| Arthropod bite | 1 (2.9) | 0 | 1 (1.7) |
| Ligament sprain | 1 (2.9) | 0 | 1 (1.7) |
| Limb injury | 1 (2.9) | 0 | 1 (1.7) |
| Muscle injury | 0 | 1 (4.2) | 1 (1.7) |
| Skin laceration | 0 | 1 (4.2) | 1 (1.7) |
| Skin wound | 1 (2.9) | 0 | 1 (1.7) |
| Wound | 1 (2.9) | 0 | 1 (1.7) |
| **Eye disorders** | 1 (2.9) | 4 (16.7) | 5 (8.6) |
| Seasonal allergy | 1 (2.9) | 3 (12.5) | 4 (6.9) |
| Central serous chorioretinopathy | 0 | 1 (4.2) | 1 (1.7) |
| **Immune system disorders** | 0 | 4 (16.7) | 4 (6.9) |
| Basedow's disease | 0 | 1 (4.2) | 1 (1.7) |
| Dermatitis contact | 0 | 1 (4.2) | 1 (1.7) |
| Food allergy | 0 | 1 (4.2) | 1 (1.7) |
| Urticaria | 0 | 1 (4.2) | 1 (1.7) |
| **Investigations** | 2 (5.9) | 2 (8.3) | 4 (6.9) |
| Aspartate aminotransferase increased | 1 (2.9) | 1 (4.2) | 2 (3.4) |
| Blood cholesterol increased | 1 (2.9) | 0 | 1 (1.7) |
| White blood cell count decreased | 0 | 1 (4.2) | 1 (1.7) |
| **Preferred Term** | **n(%)** | **n(%)** | **n(%)** |
| **Musculoskeletal and connective tissue disorders** | 3 (8.8) | 1 (4.2) | 4 (6.9) |
| Arthralgia | 1 (2.9) | 0 | 1 (1.7) |
| Back pain | 1 (2.9) | 0 | 1 (1.7) |
| Fibromyalgia | 1 (2.9) | 0 | 1 (1.7) |
| Joint injury | 0 | 1 (4.2) | 1 (1.7) |
| Rotator cuff syndrome | 1 (2.9) | 0 | 1 (1.7) |
| **Endocrine disorders** | 1 (2.9) | 2 (8.3) | 3 (5.2) |
| Diabetic ketoacidosis | 0 | 1 (4.2) | 1 (1.7) |
| Diabetic retinopathy | 1 (2.9) | 0 | 1 (1.7) |
| Insulin resistance | 0 | 1 (4.2) | 1 (1.7) |
| **Metabolism and nutrition disorders** | 1 (2.9) | 2 (8.3) | 3 (5.2) |
| Blood triglycerides increased | 1 (2.9) | 0 | 1 (1.7) |
| Hypercholesterolaemia | 0 | 1 (4.2) | 1 (1.7) |
| Hypothyroidism | 0 | 1 (4.2) | 1 (1.7) |
| **Neoplasms benign, malignant and unspecified (incl cysts and polyps)** | 1 (2.9) | 2 (8.3) | 3 (5.2) |
| Dermal cyst | 1 (2.9) | 1 (4.2) | 2 (3.4) |
| Benign breast neoplasm | 0 | 1 (4.2) | 1 (1.7) |
| **Pregnancy, puerperium and perinatal conditions** | 0 | 2 (8.3) | 2 (3.4) |
| Delivery | 0 | 1 (4.2) | 1 (1.7) |
| Pre-eclampsia | 0 | 1 (4.2) | 1 (1.7) |
| **Hepatobiliary disorders** | 0 | 1 (4.2) | 1 (1.7) |
| Hepatitis C | 0 | 1 (4.2) | 1 (1.7) |
| **Nervous system disorders** | 1 (2.9) | 0 | 1 (1.7) |
| Headache | 1 (2.9) | 0 | 1 (1.7) |
| **Psychiatric disorders** | 1 (2.9) | 0 | 1 (1.7) |
| Depression | 1 (2.9) | 0 | 1 (1.7) |
| **Reproductive system and breast disorders** | 1 (2.9) | 0 | 1 (1.7) |
| Vaginal infection | 1 (2.9) | 0 | 1 (1.7) |
| **Vascular disorders** | 0 | 1 (4.2) | 1 (1.7) |
| Hypertension | 0 | 1 (4.2) | 1 (1.7) |

| **System Organ Class** | **BCG**<br>**(N=34)** | **Placebo**<br>**(N=24)** | **Overall**<br>**(N=57)** |
| --- | --- | --- | --- |
| **Preferred Term** | **n(%)** | **n(%)** | **n(%)** |
| **Injection site reaction (Scab/Erythema)** |  |  |  |
| Total | 193 | 137 |  |
| Negative | 72 (37.3) | 136 (99.3) |  |
| Positive | 121 (62.7) | 1 (0.7) |  |

**Table 4.**

| **System Organ Class** | **BCG**<br>**(N=34)** | **Placebo**<br>**(N=24)** | **Overall**<br>**(N=58)** |
| --- | --- | --- | --- |
| **Preferred Term** | **n(%)** | **n(%)** | **n(%)** |
| **Subjects with Any Serious TEAE** | 1 (2.9) | 2 (8.3) | 3 (5.2) |
| **Endocrine disorders** | 0 | 1 (4.2) | 1 (1.7) |
| Diabetic ketoacidosis | 0 | 1 (4.2) | 1 (1.7) |
| **Gastrointestinal disorders** | 1 (2.9) | 0 | 1 (1.7) |
| Impaired gastric emptying | 1 (2.9) | 0 | 1 (1.7) |
| **Pregnancy, puerperium and perinatal conditions** | 0 | 1 (4.2) | 1 (1.7) |
| Pre-eclampsia | 0 | 1 (4.2) | 1 (1.7) |
| **Renal and urinary disorders** | 0 | 1 (4.2) | 1 (1.7) |
| Kidney infection | 0 | 1 (4.2) | 1 (1.7) |

BCG administered as an intradermal vaccine has been continuously used globally for over 100 years and excellent safety documented after over 3-4 billion doses^49^. In this trial, no moderate or serious unexpected drug related adverse effects (AE) were recorded in the BCG groups (Table 3, Table 4). The AE rate per subject over the 5 year follow-up in the BCG group (213 events for 34 subjects = 6.26 events/subject) was somewhat lower as compared to its placebo group (181 events for 24 subjects = 7.54). However, this difference was not significant (Poisson Regression Analysis p=0.124). From a clinical viewpoint, severe hypoglycemic events (i.e., requiring hospitalization, glucagon administration) and hyperglycemic events (i.e., hospitalization, diabetic ketoacidosis) were tabulated for the 5-year study period (Table 4) BCG is not known to cause systemic adverse events after administration, unlike many other vaccines or CD3 immunotherapy which can lead to symptoms like fever, chills, myalgias; no systemic AEs that were drug related were recorded in this trial.

## Discussion

In this randomized, double-blind, placebo-controlled trial, multidose BCG treatment was associated with long-term improvement in glycemic control in adults with longstanding T1D. For the prespecified primary outcome, percentage change from baseline in HbA1c, the BCG group showed significant reductions at both 5 and 8 years, and longitudinal responses differed between the BCG and placebo groups through 8 years of follow-up. A sensitivity analysis confirmed BCG specific reductions in HbA1c were robust and durable at least to Year 08. Insulin requirements declined within the BCG group only. CGM analyses further demonstrated improved glycemic profiles, including increased time in range and reduced hyperglycemia, without increased hypoglycemia. These improvements occurred without recovery of C-peptide secretion, consistent with our previous clinical studies, suggesting that the glycemic effect of BCG in longstanding T1D does not depend on restoration of pancreatic β-cell function. This clinical trial data is consistent with the mechanism of Immune Training of the diabetic host immune system to restore regulated sugar transport.

The 2-3 year delayed-onset, magnitude, and durability of the HbA1c reduction long after dosing was consistent with findings in the earlier phase 1B randomized trial^14^ and open-label trials^15,27^ of multi-dose BCG. The nuances of this trial design requires comment on the appreciation of trial design to the desired outcome. There is now an appreciation that BCG strains vary and some BCG strain have no efficacy in off target benefits; or in vitro have less potent traits^50–52^. With adult subjects receiving BCG through intradermal administration it takes 3+ years in moderately sized clinical trials to document efficacy while studies in monkeys suggest IV administration leads to rapid effects^53^. Most data supports a multi-dose regimen (six doses) rather than the single dose in adults, although data shows a single dose at birth may have durability for over 70 years. This trial uniquely uses the highly potent BCG strain (Tokyo 172)^50,52^ and it assesses durability over a much longer time horizon (up to 8 years) as compared to the typical short 1 year studies. These distinctions — multi-dosing, potent BCG strain, and long-term follow-up duration — are thought to account for maximizing the magnitude of the blood sugar effects.

The magnitude of the observed HbA1c reduction may be clinically meaningful. Mean HbA1c declined from 7.84% to 7.30% (0.54 percentage points or 6 mmol/mol) over 5 years and to 6.72% (1.12% or 12 mmol/mol) over 8 years. The extent of this drop, based on separate studies, is protective of diabetic complications^23,54–56^. More specifically, it is associated with decreased risks of cardiovascular disease and death: diabetic subjects with suboptimal glycemic control with HbA1c level over 7.5% (58.5 mmol/mol) have a strong association with increased risks of cardiovascular disease and all-cause death. In contrast HbA1c values under 7.5% are protective of the well-known diabetic complications^54,55^. Long-term studies have established a graded relationship between glycemic exposure and risk of microvascular and cardiovascular complications.

How does BCG work? Its mechanism of action appears to be, in part, metabolic. We have found that T1D patients’ lymphoid cells have a metabolic defect in glucose handling, with a relative reliance on oxidative phosphorylation and diminished capacity for aerobic glycolysis^14–16,18,19,24–26^. This defect in lymphoid metabolism is specific to T1D and is not found in type 2 diabetic subjects^15^. Lower HbA1c in BCG-treated T1Ds—in the absence of pancreatic insulin secretion—is hypothesized to result from a metabolic correction of how lymphoid cells take up and use glucose through BCG induced Immune training. In other words, BCG immunotherapy may restore lymphoid cell glucose metabolism to aerobic glycolysis, a regulated method of higher glucose uptake, glucose transport, and utilization than oxidative phosphorylation. We have previously shown, using human longitudinal^27^ FDG-PET organ mapping, that BCG treatment is associated with augmented glucose transport^18^ consistent with the timing and onset of a shift to aerobic glycolysis.

Both *Mycobacterium tuberculosis* (TB) and *Mycobacterium bovis* (BCG) demonstrate acute innate and chronic adaptive immune “training” effects. Innate immunity training changes the human immune system through systemic modulation of histones, leading to long-term opening of chromatin for gene expression (innate immune training)^28–37,39,40,42,57,58^. BCG-induced adaptive immune training effects occur over years and gradually introduce direct increases and decreases in DNA methylation, and now includes changes in the T-cells as well as monocytes^9,14,15,18,59^. In T1D, systemic acute innate immune responses increase tumor necrosis factor (TNF), which selectively kills cytotoxic autoreactive T cells according to a human trial, the original reason we started clinical trials in T1D^20–22^. Slower adaptive immune changes of lymphocytes, which occur through direct epigenetic modulations to gene-specific DNA, turn on sugar pathways, turn on the over-methylated T cell receptor of diabetes, and restore highly desirable Tregs^14,15,18,19,24,25^. Although these mechanistic studies provide a biologically plausible mechanism for the delayed glycemic effect, the present trial does not address mechanisms.

A principal limitation of BCG immunotherapy for T1D is its delayed onset of action, with clinical benefits emerging only 2-3 years after treatment initiation, although their magnitude appears to increase thereafter. In addition, this trial exclusively enrolled adults with longstanding T1D and cannot be directly extrapolated to pediatric patients. Randomized trials in pediatric patients using potent BCG strains and multidose regimens are needed to determine whether BCG can suppress autoimmunity, preserve pancreatic β-cell function and/or modify disease progression. A recent observational study reported improvements in both HbA1c and IDAA1c among patients with T1D who had received neonatal BCG vaccination compared with those who had not. As in our adult studies, these clinical findings were associated with enhanced Treg induction and increases in metabolites associated with lymphoid glycolysis^60^. Finally, prospective trials in individuals with presymptomatic T1D are needed to determine whether BCG immunotherapy can delay or prevent progression to clinical disease.

In conclusion, this lengthy phase 2 clinical trial suggests that BCG immunotherapy shows promise as a safe and durable therapeutic approach to glycemic control in longstanding T1D. Our findings demonstrate sustained glycemic improvement that persisted through 8 years of follow-up. The improvement begins 2-3 years after initial dosing and is accompanied by reduced insulin requirements and without an increase in hypoglycemia. Together, these results extend our earlier findings and support further development of BCG. Global contributions on the human benefits of off-target BCG in allergies, platform infectious disease protection, overall mortality and cancer, continue to show the synergy with BCG co-existence benefits^61–69^. BCG is an affordable therapy known to provide beneficial off-target effects, including broad protection against infectious diseases to which individuals with T1D have amongst the highest relative risks^70–72^. Thus, BCG immunotherapy may offer multiple benefits for improving long-term health in T1D.

## Data Availability

Individual subject data cannot be made available as informed consent and confidentiality agreements with subjects do not include this provision for public sharing.

## Acknowledgements

We kindly thank Dr. Miriam Davis for her proofreading and editing assistance.

## Author Contributions

Data sharing statement. At the end of the study, Drs Faustman, Kühtreiber and Zheng had full access to all the data, and take responsibility for integrity of the data, accuracy of data analysis, data interpretation. Conceptualization: Faustman. Data Curation: Kühtreiber, Advanced Clinical, Deerfield, IL; DSMB, Biologics Consulting Group, Alexandria, Virginia. Investigations: Braley, Hullavarad, Bulczynski, Dorsey, Kartsounis, Thach, Hayashi, Bradley, Mounier, Adams, Hashiguchi. Performance of Treg assays (not reported here): Tran. Formal analysis and statistics: Kühtreiber, Faustman, Zheng, Advanced Clinical, DSMB. Writing: Faustman, Kühtreiber.

## Materials & Correspondence

BCG is exclusively licensed for this trial from Japan BCG Laboratory to Massachusetts General Hospital. The other supplies are commercially available from the companies indicated.

## Methods

### Trial conduct and oversight

This trial was approved by Massachusetts General Hospital (MGH) (now Massachusetts General Brigham) Human Studies Committee with US Food and Drug Administration oversight and approval as well. This trial was an MGH institution-sponsored trial, not an investigator-sponsored clinical trial, and therefore abided by all FDA requirements for full conduct, review auditing and external oversight. The trial followed the guidelines of the World Medical Association’s Declaration of Helsinki and was registered March 7, 2014 at clinicaltrials.gov with ID NCT02081326. This was then followed by fund raising and enrollment from 2015 through 2017.

### Trial design and outcomes

This is a double-blind, randomized, placebo-controlled, phase 2 clinical trial of BCG immunotherapy (Tokyo 172 strain, Japan BCG Laboratory, Tokyo, Japan) in adults with established T1D. T1D for this study was defined as having an age of onset **<** 21 years, with autoantibodies at diagnosis, persistent hyperglycemia and insulin dependence. Eligibility and exclusion criteria are presented in Supplemental Table 2. Participants received a total of six doses of BCG or placebo: two in the first year, 1 month apart, followed by 4 annual doses. Following the Phase I double blinded clinical trial in T1D BCG treated subjects, a power analysis performed by Dr David Schoenfeld, Chief of Biostatics at Massachusetts General Hospital, recommended the Phase 2 study enroll approximately 60 T1D to reach 90% power with a 3% decline in HbA1c (SAP Version 0.0). The first unblinding of this data was at year 5 and the next unblinding of data at year 8; subjects are being followed a total of 10 years.

All study procedures and visits are outlined in Table S1. The CONSORT flow diagram outlines enrollment and randomization (Fig. 1). The primary outcome was % change in HbA1c from baseline to year 5 and year 8. Secondary outcomes included insulin dose adjusted A_1c_ (IDAA1c), exogenous insulin dose, autoantibodies and stimulated C-peptide with glucagon, a biomarker of the insulin-producing capacity of the pancreas, as direct measure of insulin cannot distinguish exogenous from endogenous sources. At the request of changing FDA guidelines, CGM data was added to this study and all monitoring extended to year 10 to show durability. All data were analyzed by an independent consulting group, Advanced Clinical Inc. (Deerfield, IL).

### Patient population

The participants were adults with established T1D, all with autoantibodies at diagnosis. Patients were eligible if they had insufficient glycemic control (HbA1c range: 6.5% -8.5%), were born in the US (to preclude prior BCG vaccinations), had never received the BCG vaccine, and were without underlying TB. Full eligibility criteria are provided (Supplemental Table 2). The trial was conducted at a single center (MGH, Boston, MA, USA), but patients were recruited and enrolled from across the continental US (Supplemental Fig. 1). Diabetes management was solely the responsibility of patients and their endocrinologists, not the trial site. To prevent bias in outcomes, no one listed as study staff performed diabetes management.

### Interventions

Subjects received 2 doses of intradermal BCG or placebo (saline) 4 weeks apart, followed by 4 yearly doses for a total of 6 doses. Each BCG dose consisted of 100 uL of BCG reconstituted in saline (0.5mg/mL). BCG and placebo syringes looked identical.

### Safety assessments

All safety data are reported for this entire trial (Table 3, 4), which covers 18 visits, all fasting. The BCG vaccine can cause erythema at the injection site, 4 weeks to 2 months afterward; no vaccine erythema exceeded the expected range as documented by a third-party blinded to study group. Study staff were blinded to the injection sites and a separate clinical nurse scored any skin reactions. Since adults with long standing diabetes have had 10-20 years of glucose monitoring and automated insulin delivery, skin reactions from the BCG were typically similar to the scarring, fibrosis and infections from monitoring device skin reactions.

### Glucagon stimulation test (GST)

At each annual visit, 1 mL intravenous glucagon (reconstituted at 1mg/mL) for the glucagon stimulation test (GST) was administered to assay β-cell function in the pancreas, causing transient nausea for 1-2 minutes. All adverse events are provided with MedDRA codes (Table 3, 4). During the GST, blood samples were taken for the preparation of serum. These samples were used for the determination of changes (AUC, Area Under the Curve, and Peak levels) of stimulated C-peptide.

Subjects were asked to adhere to a high-carbohydrate diet for the 3 days prior to the GST. The only diet requirement was to consume at least 150 grams of carbohydrates per day for the three days prior to the GST. On the morning of the GST, subjects were asked to withhold their morning dose of insulin until after they had completed the test (at which point they could eat and take insulin as they normally would). More specifically, subjects were asked to withhold rapid-acting insulin via their pump or by injection two hours before the GST, short-acting insulin six hours before the GST and long-acting insulin by injection or the pump starting midnight on the day of the GST. If, during the morning of the GST, subjects needed to take insulin due to high blood sugar, they could do so, until the range was below or equal to 225 mg/dL. Subjects who used insulin pumps could continue to receive the basal rate up to two hours prior to the GST. For subjects’ safety, we started the GST by checking and recording their blood sugars to ensure they were below or equal to 225mg/dL; if a subject’s blood sugar was greater than 225 mg/dL the morning of the GST, they were asked to do some exercise and/or bolus some short acting insulin until their blood sugar fell below or equal to 225 mg/dL and the blood sugar resumed being flat as monitored with finger sticks or through their CGM.

If the subject’s blood sugar was within the allowed range the GST proceeded, a time 0 blood sample was taken and then the IV glucagon was administered (1mg in 1 mL of saline). Subsequently, additional blood samples were collected at 2, 4, 6, 8, 10 and 20 minutes to both monitor the blood sugar and also monitor the saved sample for C-peptide levels. The area under the C-peptide curve (AUC) during the GST period was determined using the trapezoid method.

### Continuous glucose monitoring

To ensure data uniformity and at the recommendation of the FDA, Continuous Glucose Monitoring (CGM) data was collected and only used from subjects that used Dexcom monitors. In the T1D study group there were 20 BCG and 15 placebo Dexcom users. The Dexcom monitors register blood sugars every 5 minutes in a range of 40 to 400 mg/dL. We used a value of 39 to indicate a “Low” reading (<40) and a value of 401 for a “High” reading (>400). Downloaded data were stored in comma-separated-values (*.csv) files that contained date, time and blood sugar data that was recorded every 5 minutes around the clock. To synchronize CGM data for different subjects and prepare the files for analysis, a hospital assigned subject specific ID number was added to each data row in the files and the date in each row was transformed into an integer corresponding to the number of days elapsed between the start of the trial for each individual subject and the recorded CGM date. All data files were then concatenated into one large .csv file, consisting of 5,819,412 individual data points. This large file was then analyzed using custom scripts written in R versions 4.2.2 or 4.4.0 (Reference: R Core Team 2024. R: A Language and Environment for Statistical Computing. R Foundation for Statistical Computing, Vienna, Austria. https://www.R-project.org/).

For analysis, the concatenated file was read into memory and filtered for CGM data that was recorded between the start (baseline, Year 0) and the end (Year 5) of the trial for each individual subject. After this step there were 3,693,569 qualifying data points, spread over a total of 89 subjects. The CGM data was then sorted into BCG and placebo groups. For each subject in each of the 4 groups the scripts then calculated average blood glucose per 6-month period and counted the number of CGM values in the normal (70-99 mg/dL), hyperglycemic (>=180 mg/dL), and hypoglycemic (<45 mg/dL) range during each 6-month period.

In more detail, and with normoglycemia as an example, these are the steps:

- For each subject and each 6-month interval count the number of CGM timepoints that fall within the range 70-99 mg/dL. Also count the total number of CGM timepoints in that 6-month interval
- Calculate the ratio of this normoglycemia count versus the total number of CGM timepoints. This is the normoglycemia % time for that 6-month interval and for that subject.
- Calculate % change from baseline
- Average across all subjects in the BCG arm and in the placebo arm separately for each 6-month interval.

The scripts also calculated the linear regression slope across all CGM data for each individual subject separately. These various calculated results were then tabulated for further processing in GraphPad Prism Version 10 to create graphs and perform statistics. In certain cases, linear mixed effect modeling was performed using SAS. For further details, see the Results and Statistical Analysis sections.

### Statistical analysis

This 5-year trial had excellent compliance (Fig. S2). Reasons for dropping out are listed in Supplemental Table 3. The primary outcome is the percentage change in HbA1c within five and eight years after intervention in participants with T1D. Statistical methods included parametric (paired or unpaired Student’s T-test) or non-parametric (Wilcoxon or Mann-Whitney) testing to compare specific time points within or across treatment arms and also Linear Mixed Effect Modeling to compare treatment arms (BCG versus placebo) over the whole 5 or 8 year time period. Incidence rates of adverse events were analyzed using Poisson Modeling.

## Data and Code Availability

Individual subject data cannot be made available as informed consent and confidentiality agreements with subjects do not include this provision for public sharing. Data for this study has been processed in the ADaM (Analysis Data Model) and TLF generation (Tables, Figures and Listings) for auditing and verification by third party groups with the above formatting and into CDISC standards. Due to confidentiality agreements between participants and the MGH, raw individual cohort data for this trial will not be made available.

---

**Figure S1.**
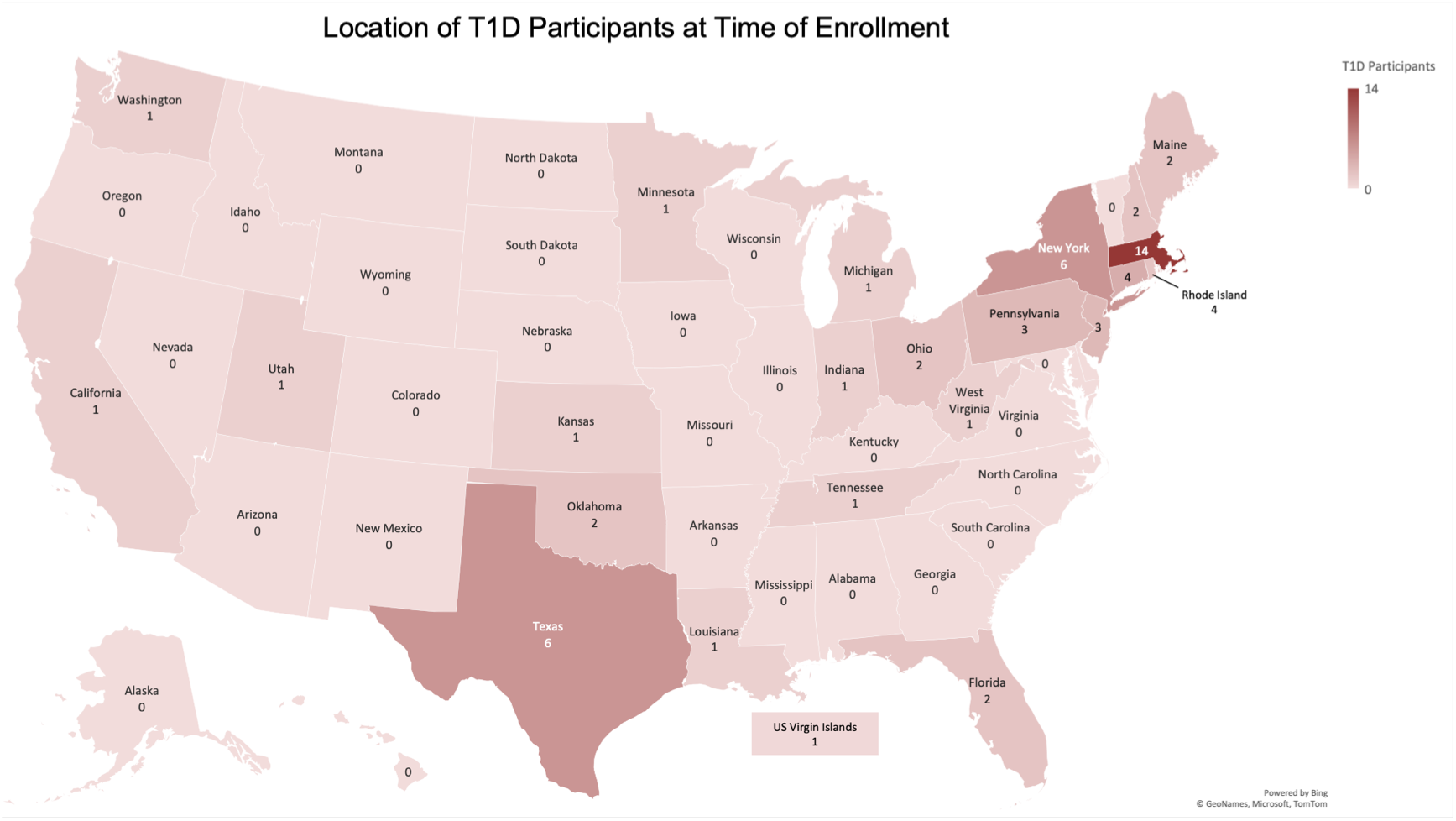
Location of T1D participants at time of enrollment This single-center trial was conducted at MGH, Boston, MA. However, patient recruitment was from across the continental United States. The map shows the number of participating T1D subjects in each state.

**Figure S2.**
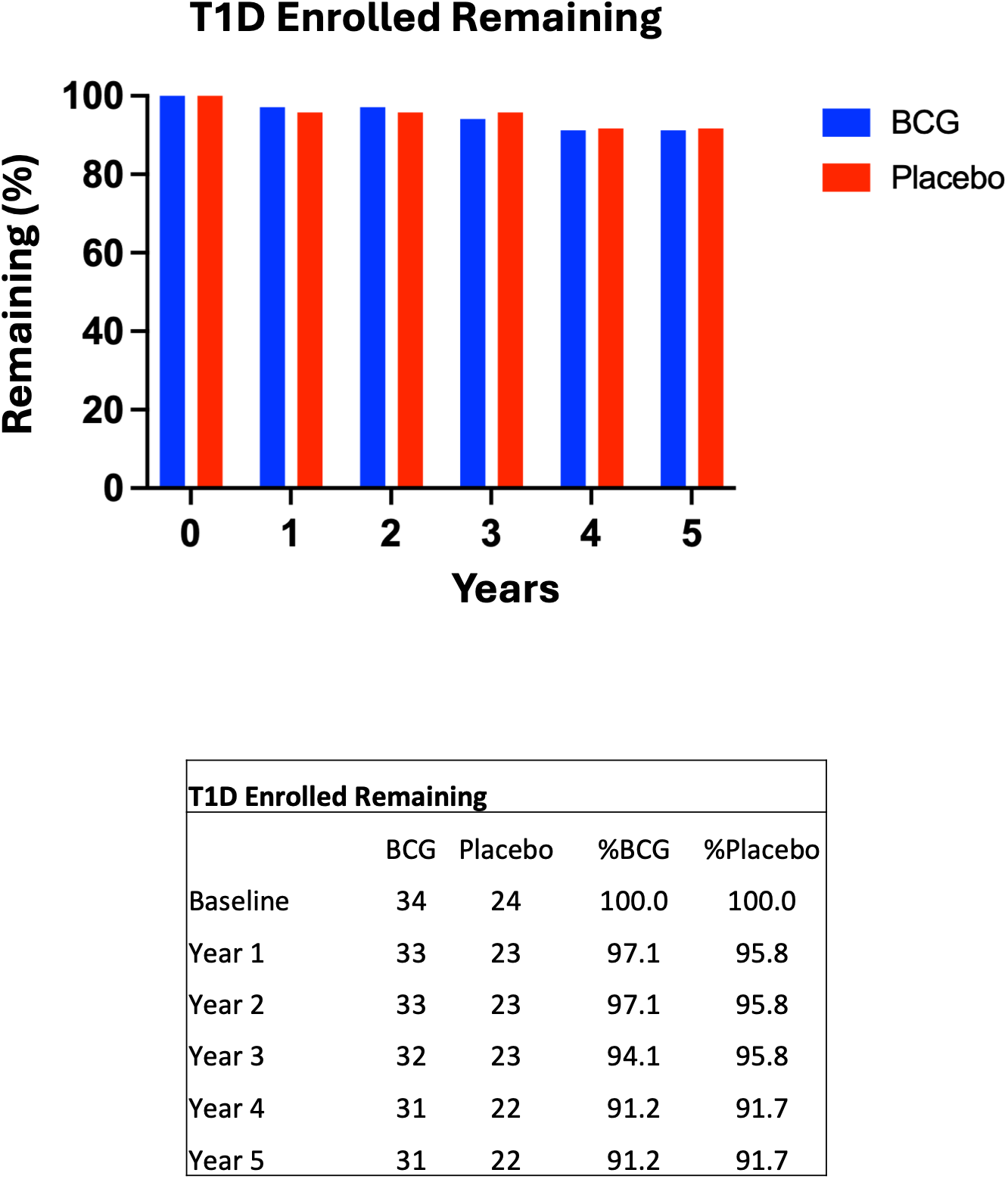
Enrolled T1D subjects remaining over time

**Figure S3.**
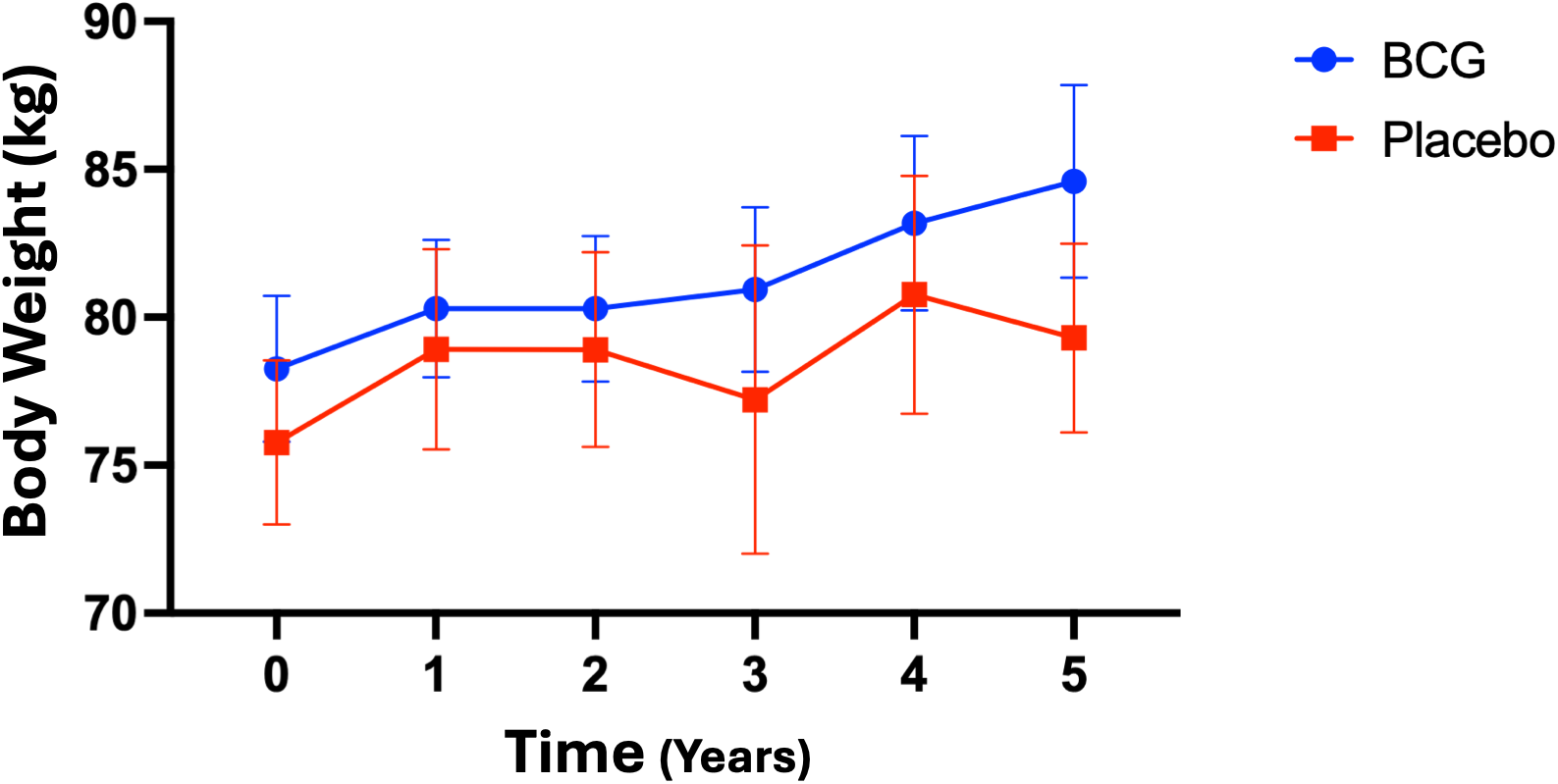
BCG has no effect on body weight There was no difference in body weight between the BCG and placebo group.

**Supplemental Table 1.**

| Overview of Study Procedures* |  |  | Follow-Up Visits and Injection Visits 2-6 |  |  |  |  |  |  |
| --- | --- | --- | --- | --- | --- | --- | --- | --- | --- |
| Visits | Screening | Randomization<br>*Injection 1* | Week 4<br>*Injection 2* | Week 8 | 6 months | Year 1<br>*Injection 3* | Year 1 +<br>4 weeks | Year 1 +<br>6 months | Year 2<br>*Injection 4* |
| Consent | X |  |  |  |  |  |  |  |  |
| CBC with<br>Differential | X | X | X |  |  | X |  |  | X |
| HIV Antibody | X | X | X |  |  | X |  |  | X |
| Comprehensive<br>Metabolic Panel | X | X | X |  |  | X |  |  | X |
| Lipid Panel | X | X | X | X | X | X | X | X | X |
| QuantiFERON-TB Gold Plus | X |  | X |  |  |  |  |  |  |
| Urine analysis | X | X | X |  |  | X |  |  | X |
| HbA1c | X | X | X | X | X | X | X | X | X |
| Proteomics | X | X | X | X | X | X | X | X | X |
| HCG (urine) |  | X | X |  |  | X |  |  | X |
| **BCG or Placebo**<br>**Injection** |  | **X** | **X** |  |  | **X** |  |  | **X** |
| Insulin, Proteins,<br>GAD,<br>Metabolomics | X | X | X | X | X | X | X | X | X |
| Autoreactive T<br>cells/Tregs | X | X | X | X | X | X | X | X | X |
| Serum C-peptide | X | X | X | X | X | X | X | X | X |
| Immune Profile<br>Assay (only<br>drawn once, but<br>can occur at any<br>visit) |  | X | X | X | X | X | X | X | X |
| Vaccination Site<br>Monitoring |  |  | X | X |  |  | X |  |  |
| GST |  | X | X |  |  | X |  |  | X |
| Visit Sheet<br>(survey of<br>medications,<br>insulin delivery<br>method and<br>safety questions) | X | X | X | X | X | X | X | X | X |
| Review of<br>complete MR for<br>IDDM<br>complications,<br>HbA1c | X |  |  |  |  |  |  |  |  |

\*Continuous Glucose Monitor data was downloaded on a bi-monthly basis for patients who provided optional consent and who use a Dexcom device.

| Follow-Up Visits and Injections 2-6, Continued |  |  |  |  |  |  |  |  |  |
| --- | --- | --- | --- | --- | --- | --- | --- | --- | --- |
| Visits | Year 2 +<br>4 weeks | Year 2 +<br>6 months | Year 3<br>*Injection 5* | Year 3 +<br>4 weeks | Year 3 +<br>6 months | Year 4<br>*Injection 6* | Year 4 +<br>4 weeks | Year 4 +<br>6 months | Year 5 |
| Consent |  |  |  |  |  |  |  |  |  |
| CBC with Differential |  |  | X |  |  | X |  |  | X |
| HIV Antibody |  |  | X |  |  | X |  |  | X |
| Comprehensive Metabolic Panel |  |  | X |  |  | X |  |  | X |
| Lipid Panel |  | X | X |  | X | X |  | X | X |
| QuantiFERON-TB Gold Plus |  |  |  |  |  |  |  |  |  |
| Urine analysis |  |  | X |  |  | X |  |  | X |
| HbA1c |  | X | X |  | X | X |  | X | X |
| Proteomics |  | X | X |  | X | X |  | X | X |
| HCG (urine) |  |  | X |  |  | X |  |  |  |
| **BCG or Placebo Injection** |  |  | X |  |  | X |  |  |  |
| Insulin, Proteins, GAD, Metabolomics |  | X | X |  | X | X |  | X | X |
| Autoreactive T cells/Tregs |  | X | X |  | X | X |  | X | X |
| Serum C-peptide |  | X | X |  | X | X |  | X | X |
| Immune Profile Assay (only drawn once, but can occur at any visit) |  | X | X |  | X | X |  | X | X |
| Vaccination Site Monitoring | X |  |  | X |  |  | X |  |  |
| GST |  |  | X |  |  | X |  |  | X |
| Visit Sheet (survey of medications, insulin delivery method and safety questions) | X | X | X | X | X | X | X | X | X |
| Review of complete MR for IDDM complications, HbA1c |  |  |  |  |  |  |  |  |  |

\*Continuous Glucose Monitor data was downloaded on a bi-monthly basis for patients who provided optional consent and who use a Dexcom device (n = 89)

**Supplemental Table 2.**
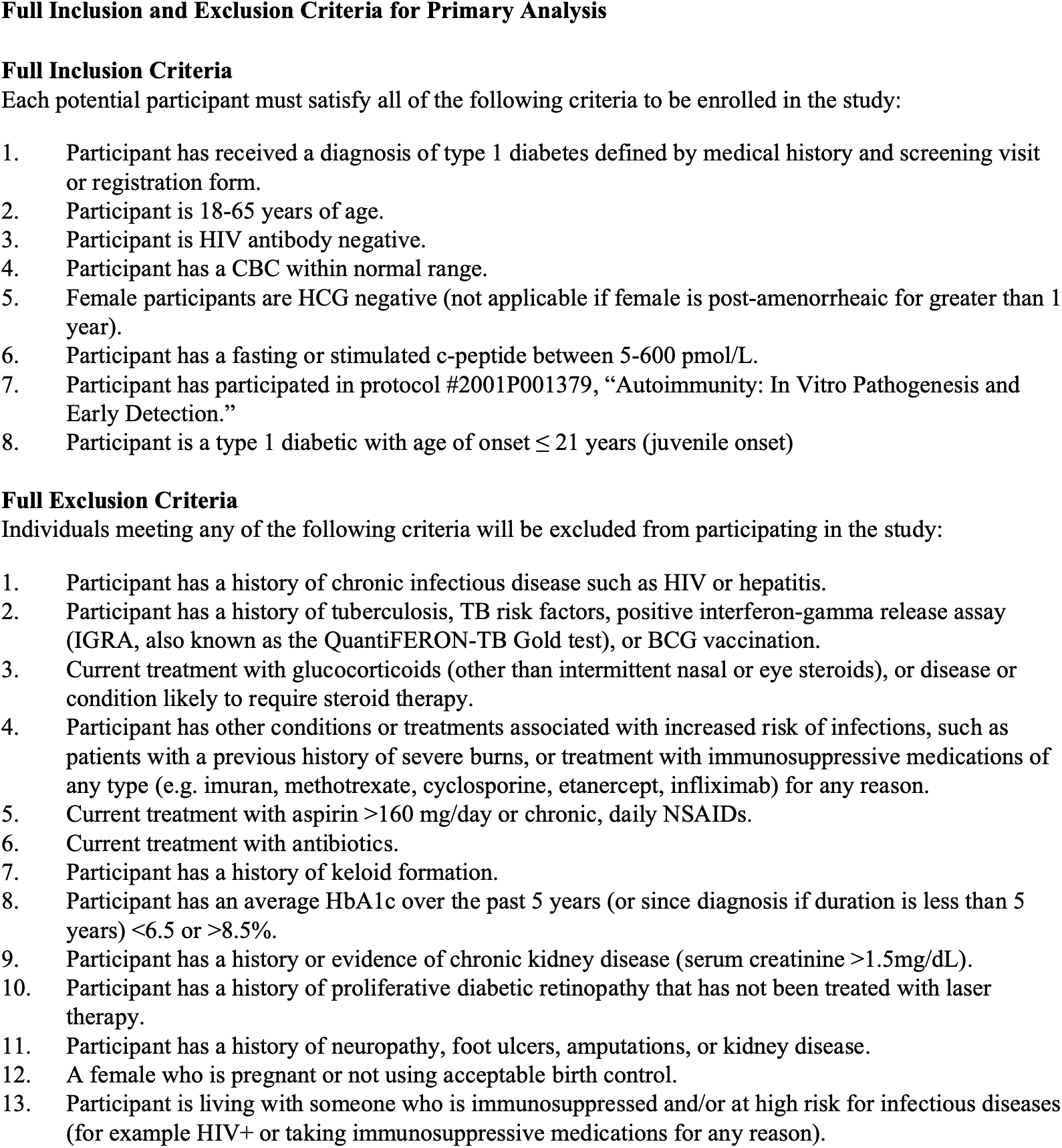

**Supplemental Table 3.**
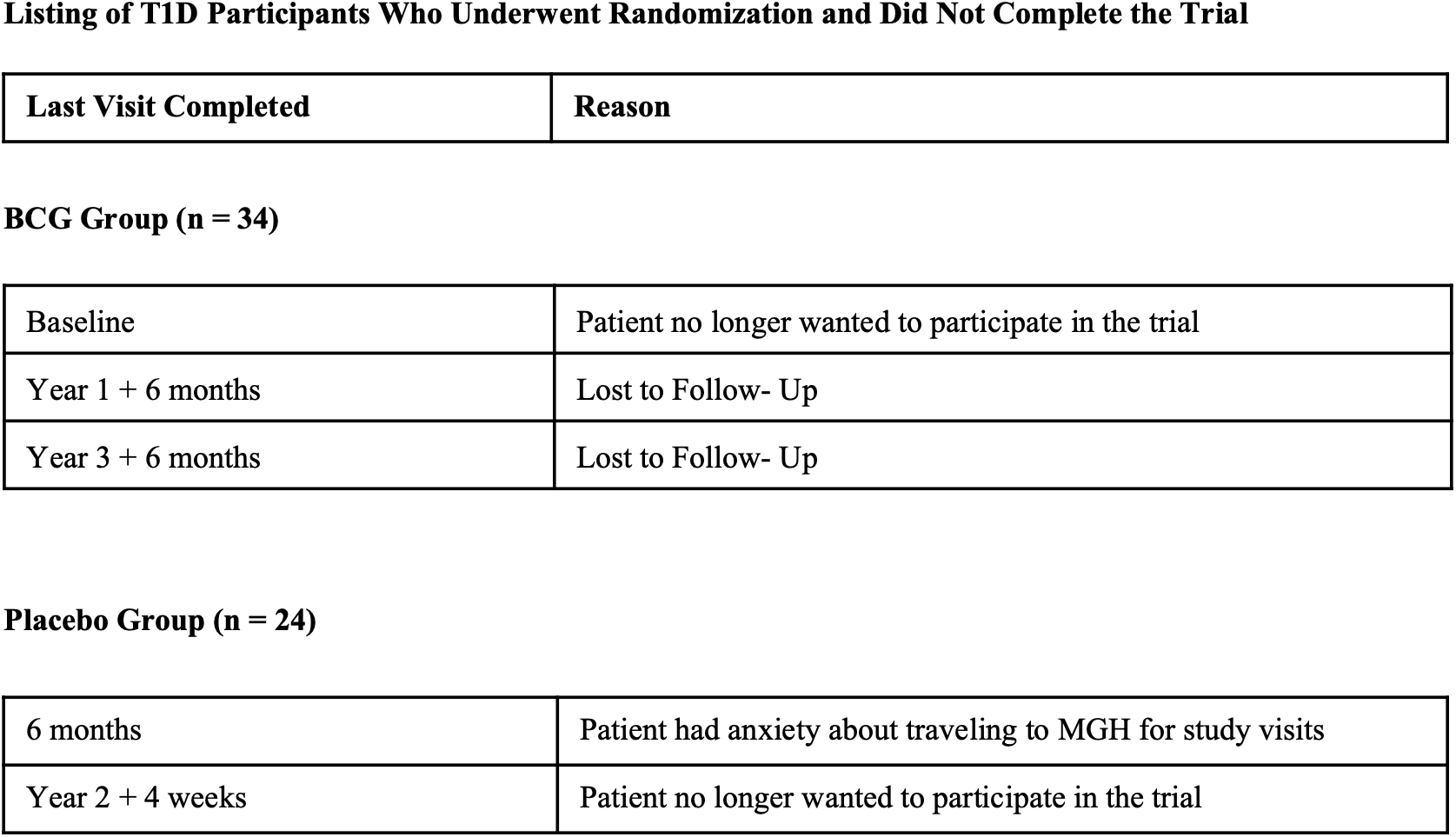

| **Last Visit Completed** | **Reason** |
| --- | --- |
| **BCG Group (n = 34)** |  |
| Baseline | Patient no longer wanted to participate in the trial |
| Year 1 + 6 months | Lost to Follow- Up |
| Year 3 + 6 months | Lost to Follow- Up |
| **Placebo Group (n = 24)** |  |
| 6 months | Patient had anxiety about traveling to MGH for study visits |
| Year 2 + 4 weeks | Patient no longer wanted to participate in the trial |

